# A phenome-wide association study of CNVs genotyped from genome sequencing read depth in the UK Biobank

**DOI:** 10.1101/2025.08.05.25333035

**Authors:** Paras Garg, Bharati Jadhav, Mariya Shadrina, Alejandro Martin-Trujillo, Andrew J. Sharp

**Author notes:** Address for correspondence: Andrew J. Sharp, Department of Genetics and Genomic Sciences, Icahn School of Medicine at Mount, Hess Center for Science and Medicine, 1470 Madison Avenue, Room 8-116, Box 1498, New York, NY 10029 USA.

## Abstract

Using genome sequencing read depth, we genotyped CNVs within the UK Biobank and performed PheWAS, identifying 501 CNVs associated with 1,537 traits. We detected signals with mosaic, recurrent and multiallelic CNVs that are difficult to genotype using other methods, such as a coding repeat within mucin 1 associated with stomach/duodenal polyps and copy number of salivary amylase genes associated with denture use. We also identified intergenic CNVs with strong effects on traits known to be regulated by nearby genes. For example, carriers of a rare non-coding deletion ∼100-kb upstream of *MC4R*, coding mutations in which are the most common cause of monogenic obesity, were an average of ∼14 kg heavier than controls. Our study provides a detailed map of functional CNVs, including complex loci that are recalcitrant to other methods, providing numerous insights into their effects on human traits.

## Introduction

Genome-wide association studies typically rely on genotyping large numbers of single nucleotide variants (SNVs) to identify disease associations. Contrastingly, relatively few association studies have analyzed copy number variants (CNVs), despite their prevalence in the genome and known links with disease^1,2,3^.

While several phenome-wide association studies (PheWAS) have utilized data from microarrays to genotype CNVs, such studies are hampered by low resolution and poor coverage in complex regions of the genome^4,5^. Alternative approaches using imputation perform poorly for rare and recurrent events^6^, while methods that utilize exome sequencing are limited to analysis of coding regions^7,8^. Improved genotyping of CNVs can be performed using genome sequencing (GS) data. However, due to their structural complexity, with highly variable sizes, breakpoints and multiple alleles that can have differing degrees of overlap, there are considerable challenges utilizing standard CNV genotypes in association studies^9^.

To overcome these limitations, we developed a read depth-based approach from whole genome sequencing data that allows accurate and scalable genotyping of copy number (CN) across the entire allelic spectrum and which produces CNV genotypes in a format that can be easily utilized in PheWAS. With this approach, we generated tiled CNV maps at 5-kb resolution throughout the genome in >490,000 individuals from the UK Biobank (UKB). Using these genotypes, we performed CNV PheWAS of 13,215 traits, testing three different inheritance models that reflect different underlying mechanisms by which CNV can alter disease risk. Our results significantly extend current knowledge of the effects of CNVs on human traits.

## Results

### Sequencing read depth provides highly accurate genotyping of CNVs at 5kb resolution

We developed an efficient pipeline based on read depth from GS to derive CN estimates in the UKB cohort. After data normalization and quality filtering, we used an automated clustering approach to convert quantitative CN estimates to integer CN genotypes for contiguous non-overlapping 5-kb bins throughout the genome per individual. This pipeline yielded a CNV call set in all UKB samples that we used to perform a CNV PheWAS, summarized in Figure 1. Example results of our CN genotyping method for the highly variable *CYP2A6* locus are shown in Figure 1b, with additional highly variable loci shown in Supplementary Figure 1.

**Figure 1.**
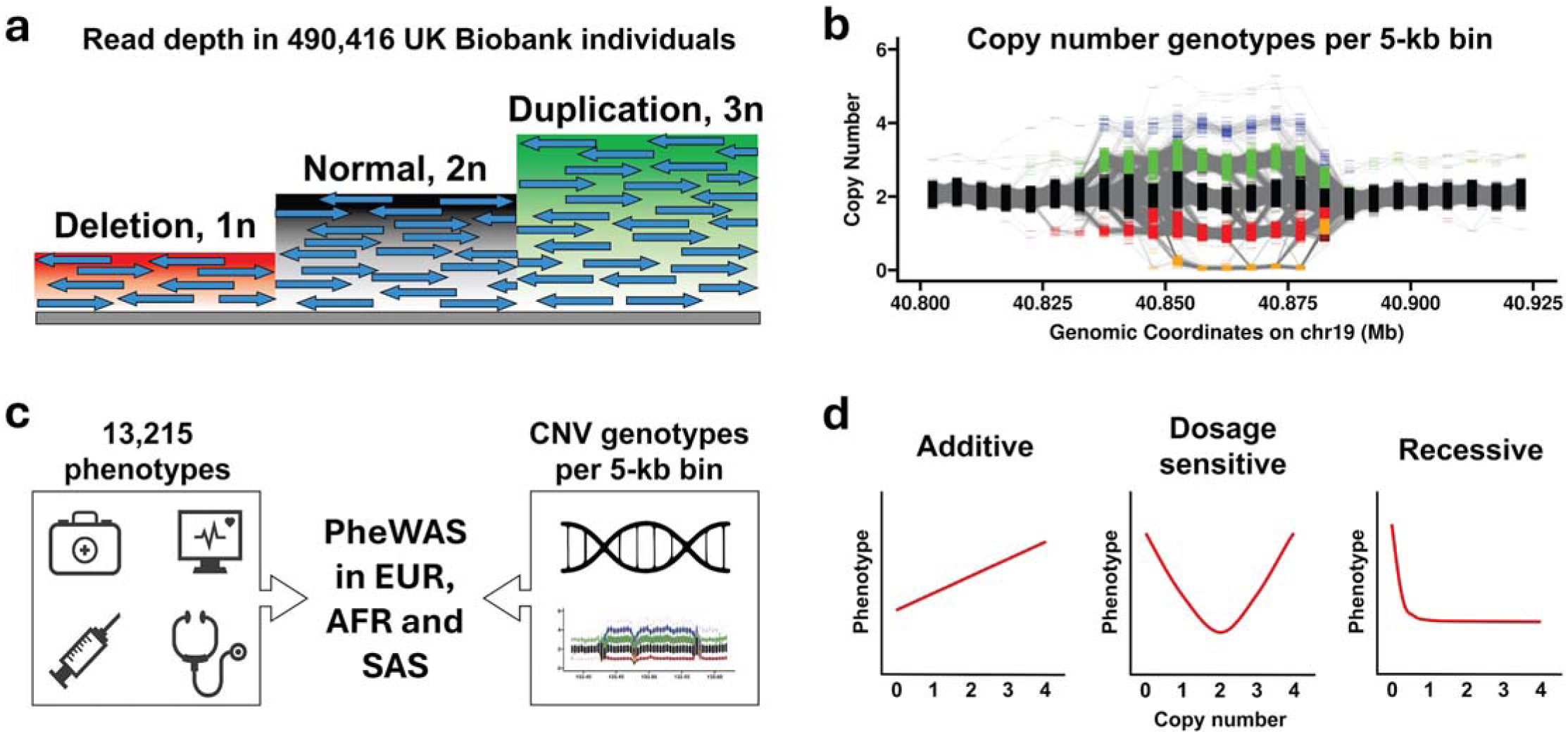
Graphical overview of the study. **(a)** We used normalized read depth from GS to estimate DNA CN in the UKB cohort. **(b)** Read depth measurements were summarized into contiguous non-overlapping 5-kb bins tiled throughout the genome and converted to CN genotypes using an automated algorithm. We show CN genotypes at the *CYP2A6* locus at 19q13.2 obtained from 405,362 unrelated individuals of European ancestry. Each color represents a different CN cluster identified by automated genotyping, with grey lines joining CN states at adjacent 5-kb bins in each individual, allowing CNV breakpoints to be visualized. **(c)** We utilized data for 13,215 phenotypes and CNV genotypes per 5-kb bin tiled across the genome to perform PheWAS in cohorts of unrelated European, African and South Asian ancestry, **(d)** testing three models representing different biological mechanisms by which change in genomic CN can lead to a phenotype (see Methods). Note that for each model, the direction of effect may be reversed to that shown.

As an initial quality check of our data, we utilized 146 pairs of monozygotic (MZ) twins of European ancestry within the UKB. Given that both members of MZ twin pairs are derived from a single embryo, barring any somatic variants, MZ twin pairs are expected to be genetically identical. Thus, genotype concordance within MZ twin pairs provides a simple benchmark to assess genotyping accuracy. Considering all 5-kb windows in the genome that we utilized for PheWAS, CN genotypes showed an average intra-MZ twin pair concordance of 99.10% (range 98.96 - 99.16%), with the vast majority of discordancies representing differences of a single copy at highly variable multiallelic loci. After excluding these multiallelic loci (5-kb bins with optimal CN model ≥ 2, representing ∼3.5% of the genome), mean intra-MZ twin pair genotyping concordance increased to 99.98% (range 99.94 - 99.99%), corresponding to an average of one genotyping error every ∼55-Mb per individual. These observations indicate the high accuracy of our read depth and automated CN assignment approach for genotyping CNVs, which was confirmed by visual inspection of genotype clusters at CNV loci.

Consistent with deletions typically being more damaging than duplications^10^ and the effects of purifying selection^11^, CNV allele frequencies varied by type, gene overlap and gene intolerance to loss-of-function SNVs (Supplementary Figure 2). For example, intergenic duplications were, on average, 3.8-fold more common than deletions that encompassed exons of constrained genes (LOEUF < 0.6)^12^.

### PheWAS identifies hundreds of CNVs associated with human traits

We extracted data for 13,215 quantitative, categorical and binary traits measured in UKB participants. Using available phenotype and genome-wide CN data, we performed PheWAS using 405,362 unrelated, quality-filtered UKB participants of European ancestry. For each 5-kb bin tiled across the genome that showed sufficient variation to be used in PheWAS and passed our quality controls, we tested three models that reflect different underlying mechanisms by which CNVs can modify traits: an additive model, a dosage sensitive model and a recessive model (Figure 1d, see Methods).

After applying a multiple testing correction based on the number of independent traits and CNVs tested, we identified 5,626 5-kb bins where CNV of the region was significantly associated with one or more traits (Bonferroni p < 3.9 x 10^-10^). However, as many CNVs are large and encompass multiple contiguous 5-kb bins, we reduced this to a set of 501 unique CNVs (482 autosomal and 19 X-linked), representing 4,477 unique pairwise CNV:trait associations. Of these, 3,411 (76%) were identified using the additive association model, 922 (21%) showed the strongest signal using a dosage sensitive model and 144 (3%) using a recessive model. A Manhattan plot showing genome- and phenome-wide results is shown in Figure 2, with example results for three individual loci shown in Figure 3. Full results for autosomes and chromosome X are shown in Supplementary Tables 1 and 2, respectively. In addition, we highlight 60 CNV:trait associations of particular biological interest in Table 1. In support of our methodology, we replicated dozens of previously reported gene:phenotype associations and also identified many other signals where the associated phenotype has a highly plausible link with the known function of gene(s) that either overlap or are adjacent to the CNV.

**Figure 2.**
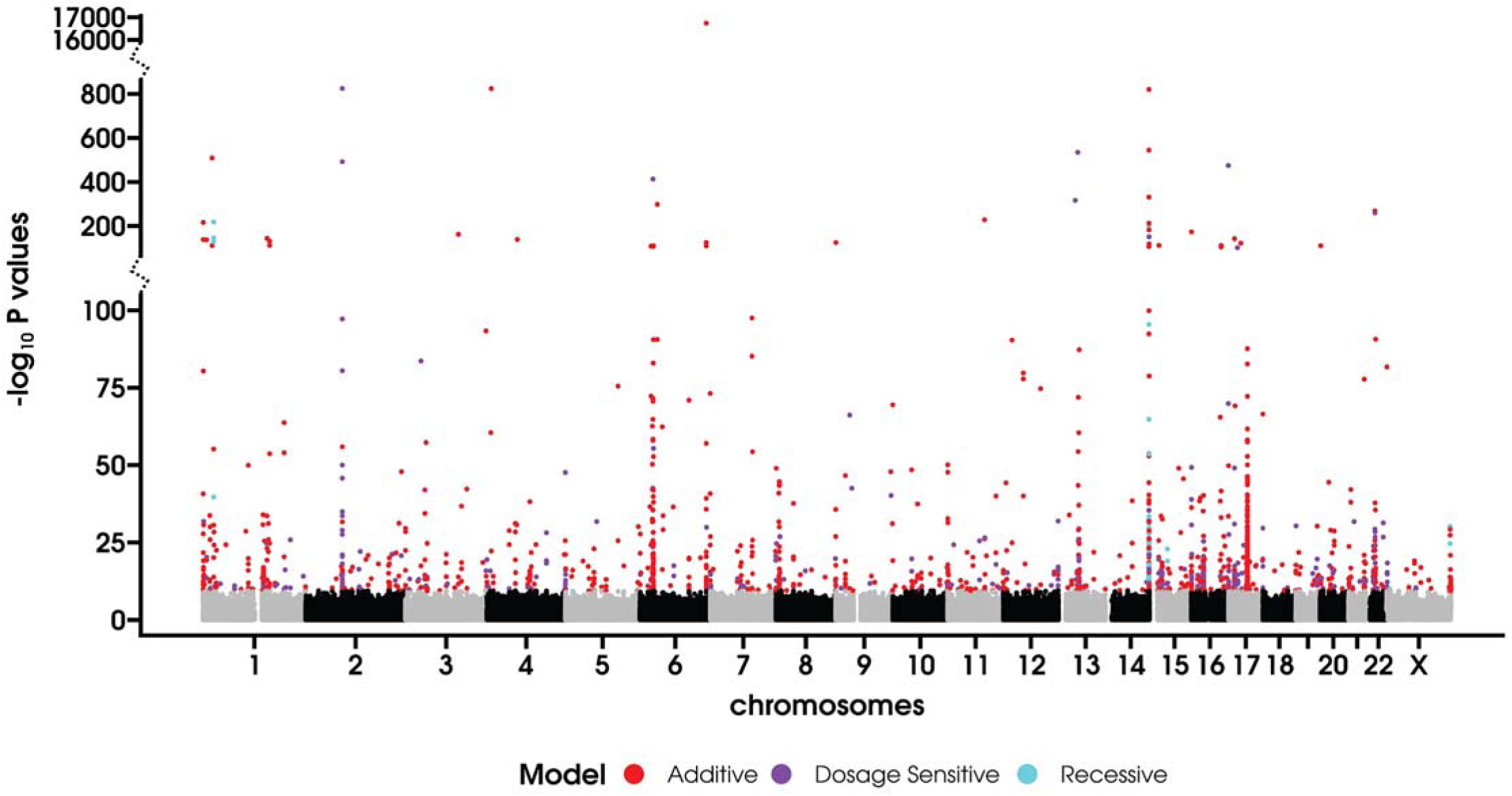
Manhattan plot showing genome-wide PheWAS results in 405,362 Europeans. Plot shows all results obtained using three association models (additive, dosage sensitive and recessive) on chromosomes 1-22 and chromosome X. To avoid redundancy, for each CNV:trait pair, we only plot results for the single 5-kb bin and model that showed the peak p-value. Color of each point indicates significant associations (Bonferroni p < 0.05) based on the association model that yielded the peak p-value: red for additive, purple for dosage sensitive and cyan for recessive. Note the use of a discontinuous y-axis to accommodate signals with extreme p-values.

**Figure 3.**
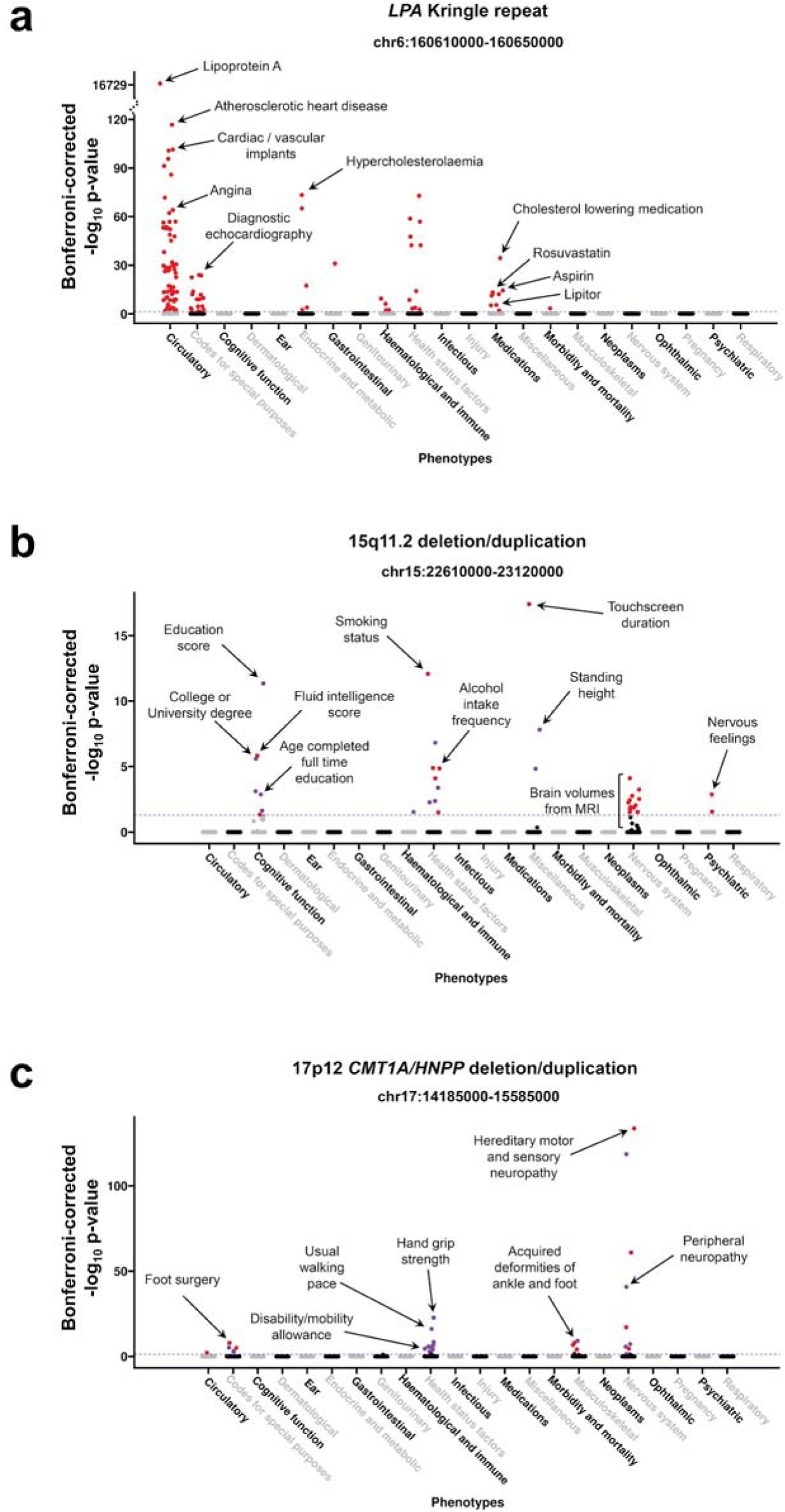
PheWAS results at three CNV loci in 405,362 Europeans. **(a)** The Kringle coding repeat within *LPA*. **(b)** The recurrent 15q11.2 deletion/duplication. **(c)** The recurrent 17p12 CMT1A*/HNPP* deletion/duplication. Coordinates above each plot indicate the coordinates of the CNV region where signals were observed. Traits are grouped into physiological categories (x-axis), with select traits individually labeled. Y-axis shows Bonferroni-corrected p-values, with the dashed grey horizontal line showing the significance threshold of p < 3.01 x 10^-10^ (Bonferroni p < 0.05) with signals exceeding this shown as colored points. Color of each point indicates the association model that yielded the peak p-value: red for the additive model and purple for the dosage sensitive model. In **(a)**, note the discontinuous y-axis used in the upper plot due to the very strong signal observed for Lipoprotein A levels.

**Table 1.**
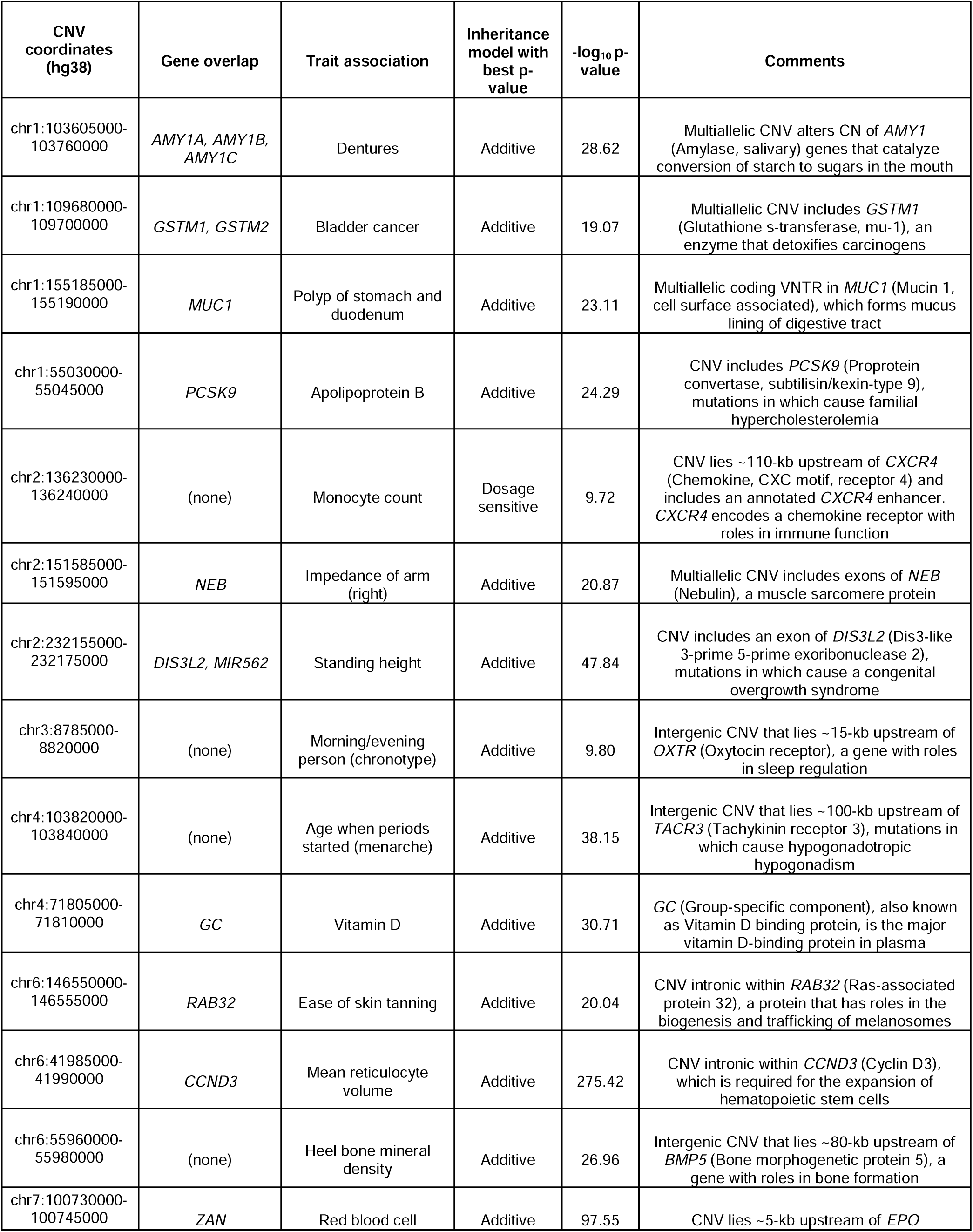
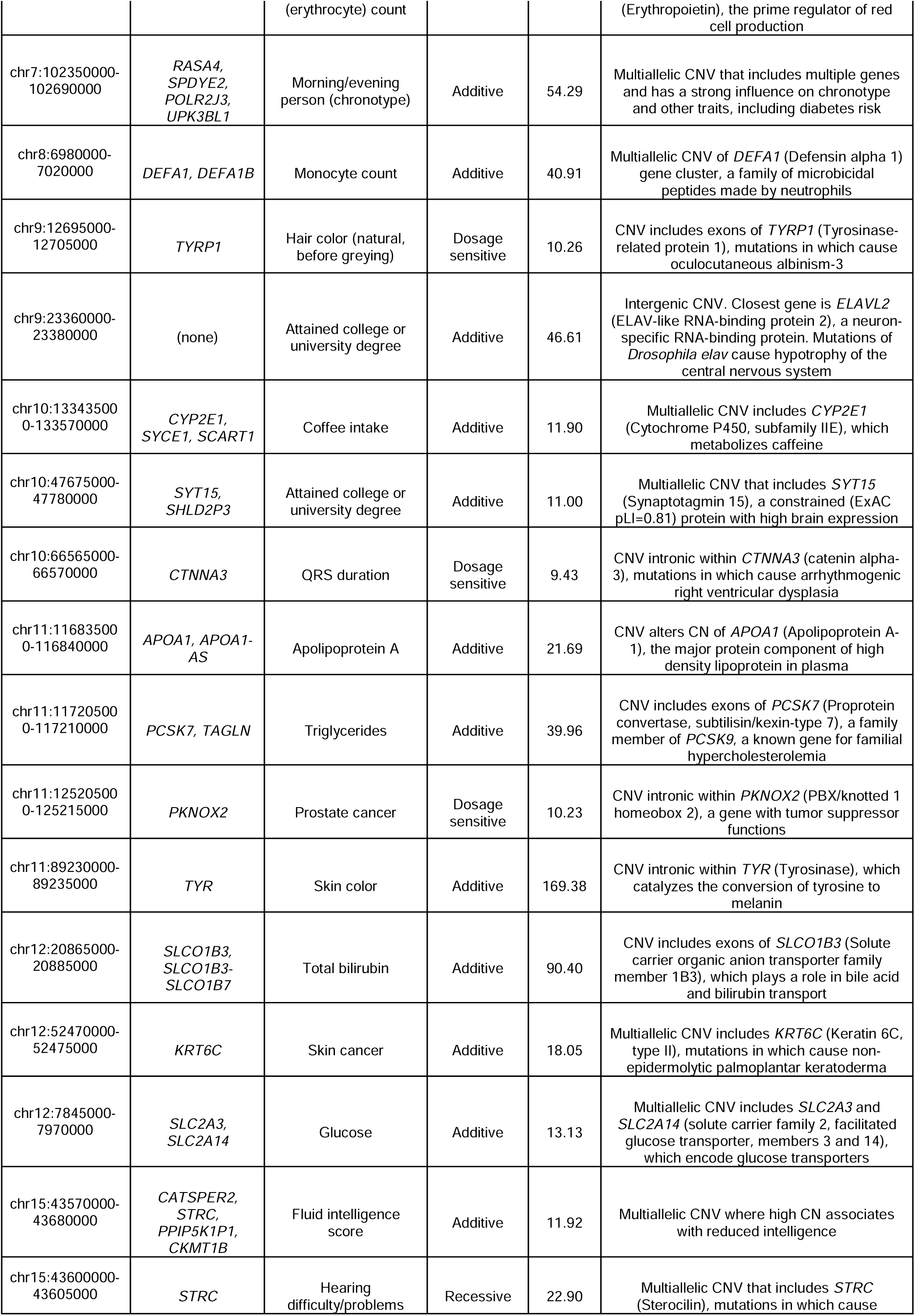
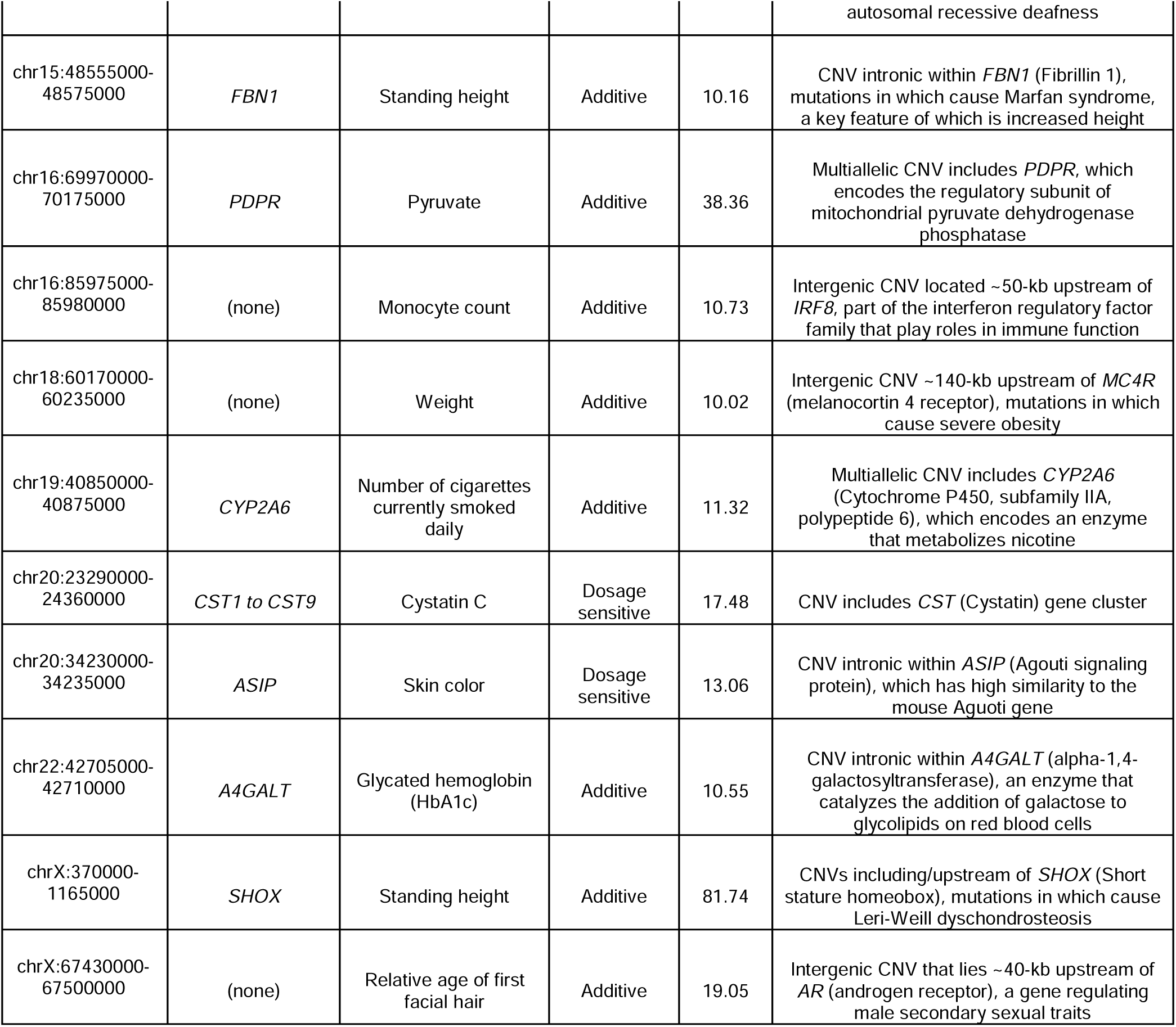
Notable CNV associations identified in Europeans.

### Multiallelic variants

Large multiallelic and repetitive regions can be particularly challenging to genotype using many technologies, leading to considerable uncertainty and controversy in their role in regulating human traits^13,14^. Using read depth, we were able to generate robust estimates of diploid CN at these loci. In addition to confirming previously reported associations such as CN of the Kringle coding repeat within *LPA* with lipoprotein A levels (Figure 3a, p = 4 x 10^-16,737^) and risk of atherosclerotic heart disease (p = 1 x 10^-125^)^15^, *FCGR3B* and serum protein levels (p = 1.3 x 10^-131^)^16^, *DEFA1* (alpha defensin) genes and monocyte count (p = 1.2 x 10^-41^)^17^ and *RASA4* with chronotype (p = 5.2 x 10^-55^) and risk of diabetes (p = 1.3 x 10^-15^)^17^, we also identified several novel associations attributable to CNV of large repeats. For example, we observed a strong positive association between CN of the highly polymorphic *AMY1* (salivary amylase) and denture use (p = 2.4 x 10^-29^) (Figure 4a), which we hypothesize is likely attributable to the role of salivary amylase in converting starch to sugars in the mouth. Notably, in contrast to prior reports that used techniques such as qPCR to genotype *AMY1* CN^18,19^, we observed no evidence of association between *AMY1* CN and obesity, diabetes or related metabolic traits (all p > 0.01), despite our very large sample size that was well powered to detect such effects.

**Figure 4.**
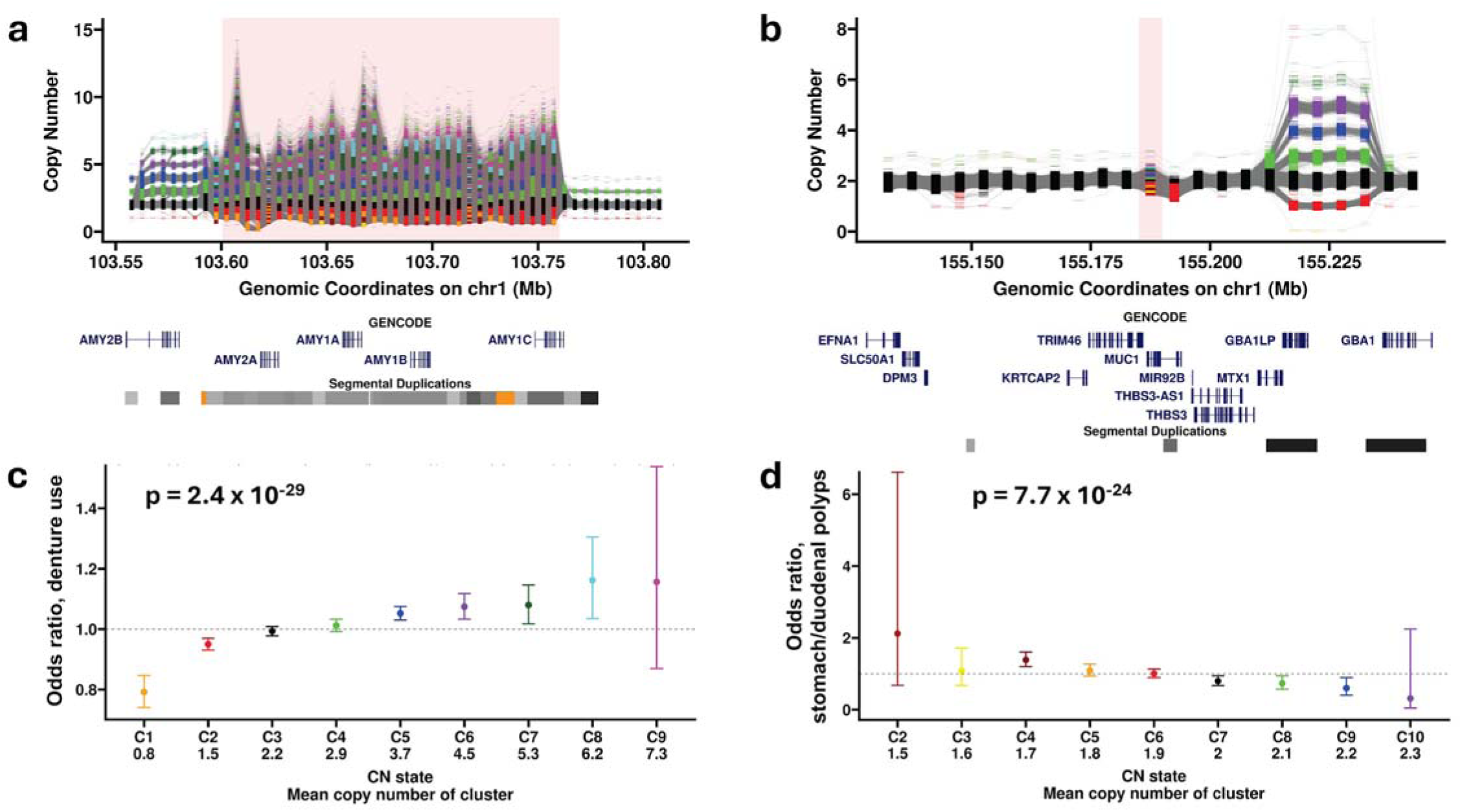
Trait associations with multiallelic CNVs. **(a** and **c)** Copy number of salivary amylase (*AMY1*) genes is positively associated with denture use. **(b** and **d)** Copy number of a coding repeat within mucin 1 (*MUC1*) is negatively associated with risk of stomach and duodenal polyps. **(a** and **b)** show CN estimates per 5-kb bin across each locus, with red shading indicating the extent of the CNV region that was significantly associated with the trait. Colors represent CN clusters identified by automated genotyping, with grey lines joining CN calls at adjacent 5-kb bins for each individual, allowing CNV breakpoints to be visualized. Below each plot are shown annotations of genes and segmental duplications. In **(c** and **d)**, colors of each bar correspond to CN clusters identified at each CNV, as shown in **(a** and **b)**, with vertical bars indicating 95% confidence intervals of odds ratios. In **(c)**, odds ratios and p-value are for the 5-kb bin chr1:103,755,000-103,760,000. Odds ratios are only shown for CN bins with ≥100 individuals.

Another example was the negative association between CN of a large coding repeat within *MUC1* (mucin 1) and several traits, including risk of stomach/duodenal polyps (p = 7.7 x 10^-24^) (Figure 4b). *MUC1* is a transmembrane glycoprotein that forms the mucous lining of the digestive tract and other organs, providing a protective barrier against infection^20^. As each copy of the coding repeat encodes one unit of the extracellular domain, shorter *MUC1* repeat alleles likely result in production of mucus with attenuated barrier function. These results complement prior targeted studies that suggested smaller *MUC1* repeat alleles are more prevalent in individuals with gastric cancer^21,22^. We also observed a positive association between a multiallelic CNV of *KRT6C* (keratin 6C) and risk of malignant skin neoplasms (p = 8.8 x 10^-19^), a positive association between CN of a multiallelic region that includes several exons of *NEB* (nebulin, a muscle sarcomere protein), and measures of muscle mass from bioelectrical impedance analysis (p = 1.3 x 10^-21^) (Supplementary Figure 3), a positive association between a multiallelic CNV overlapping the non-coding RNA *LINC01001* and risk of both uterine fibroids (p = 9.4 x 10^-12^) and prostate hyperplasia (p = 1.9 x 10^-11^), and a positive association between CN of a GC-rich VNTR intronic within *CASP8* and risk of malignant skin neoplasms (p = 5.6 x 10^-13^) (Supplementary Figure 4).

### Reciprocal recurrent gain/loss CNVs

Large recurrent reciprocal deletions and duplications underlie several common genetic syndromes and often exhibit pleiotropic effects, with several associated with increased risk for a variety of congenital malformations, neurodevelopmental and psychiatric traits^1^. Consistent with prior studies^23,24^, we identified dozens of traits indicative of increased morbidities associated with some of the most common recurrent genomic disorders, including CNVs at 1q21.1, 15q11.2, 15q13.3, 16p13.11, 16p11.2, 17p12, 17q21.31 and 22q11.21 (Figure 3, Supplementary Table 1). In addition, the relative effect sizes and allele frequencies we observed at these recurrent CNVs supports the notion that deletions of dosage sensitive genes typically have more damaging effects than reciprocal duplications^25,26^ (Supplementary Figure 5).

Reciprocal deletion/duplication CNVs occur as a result of meiotic non-allelic homologous recombination between highly identical flanking repeats and often exhibit significantly elevated mutation rates^27^. Loci prone to these recurrent rearrangements are typically characterized by multiple unrelated individuals in the population who carry reciprocal gain and loss CNVs with identical breakpoints that map to paired flanking repeat elements. We observed numerous examples of loci consistent with this expectation. For example, we identified a rare (MAF ∼0.1%) ∼20-kb reciprocal deletion/duplication CNV within *DIS3L2* that has a strong influence on standing height and measures of body mass and which appeared to be mediated by a pair of flanking L1 elements. This CNV is associated with an average of ∼5 cm increase in standing height with each additional copy (p = 1.4 x 10^-48^) and causes an in-frame gain/loss of exon 9 of *DIS3L2*, biallelic coding mutations in which cause a recessive overgrowth syndrome^28^ (Supplementary Figure 6). A second example was a ∼5-kb reciprocal deletion/duplication event that has paired L1P elements at each breakpoint and overlaps the final exon of the non-coding transcript *LOC100506444*, the duplication allele (MAF ∼0.005) of which associated with a 3.4-fold increased risk of uterine fibroids (p = 4.8 x 10^-25^) (Supplementary Figure 7).

We also observed several larger regions with reciprocal gain and loss CNVs where paired segmental duplications mapped to both breakpoints, resulting in at least five different CN states of the intervening region segregating in the population. These include the *CFHR* locus where CN positively associated with risk of macular degeneration (p = 6.9 x 10^-12^) (Supplementary Figure 1a), the *PDPR* locus where CN negatively associated with serum pyruvate (p = 4.3 x 10^-39^) and alanine (p = 3.7 x 10^-66^) levels (Supplementary Figure 1b), a region containing the hexose transporters *SLC2A3* and *SLC2A14* where CNV negatively associated with both serum glucose levels (p = 7.4 x 10^-14^) and age of menarche (p = 4 x 10^-23^) (Supplementary Figure 8) and the *CYP2E1* locus where CN showed reciprocal associations with serum acetone levels (p = 8.3 x 10^-51^) and coffee intake (p = 1.2 x 10^-12^) (Supplementary Figure 9).

An ∼200kb region in 15q15.3 exhibiting similar multiallelic gain and loss was of particular interest. This CNV contains seven RefSeq genes, including *STRC* and *CATSPER2*, mutations of which cause recessive sensorineural deafness and male infertility, respectively^29^. Consistent with this, under our recessive model, we detected homozygous deletions of this region associated with an ∼21-fold increased risk of hearing aid use (p = 9.5 x 10^-20^). However, we also identified a significant inverse dosage relationship between CN of this region with fluid intelligence (p = 1.2 x 10^-12^), which is a composite measure of cognitive function based on multiple tests of memory, language, numerical administered to UKB participants (Figure 5). The magnitude of this effect on fluid intelligence was considerable, with carriers of four copies versus zero copies of this locus showing a mean reduction in fluid intelligence scores that was comparable to the negative effect seen in carriers of the recurrent 16p11.2 deletion^30^. Of note, this region is unusual amongst reciprocal deletion/duplication loci in that the frequency of the deletion allele, which is associated with increased fluid intelligence, is approximately double that of the reciprocal duplication, even though these deletions cause male infertility when present in homozygosity.

**Figure 5.**
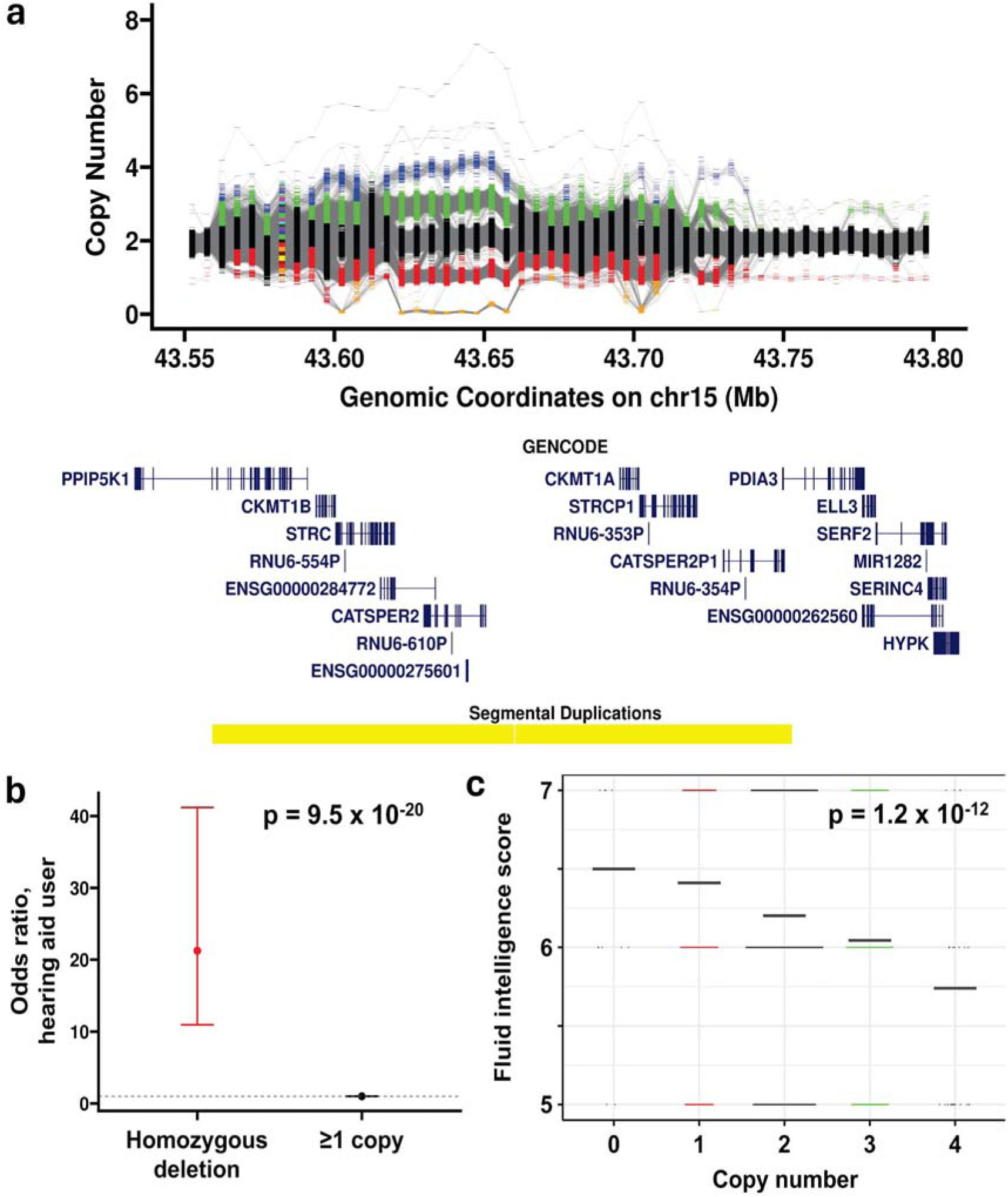
A multiallelic CNV in 15q15.3 shows opposing effects on hearing and fluid intelligence. **(a)** Homozygous deletions of an ∼200-kb region containing stereocilin (*STRC*) are associated with **(b)** a ∼21-fold increased risk of hearing aid use (p = 9.5 x 10^-20^). **(c)** The same region shows a negative dose-dependent association between copy number and fluid intelligence (p = 1.2 x 10^-12^). In **(b)**, vertical bars indicate 95% confidence intervals of odds ratios. In **(c)**, thick horizontal black bars show the mean fluid intelligence score per CN bin, with data shown only for CN bins with ≥20 individuals. Odds ratios and p-values are shown for the 5-kb bin with the peak p-value for hearing aid use (chr15:43,600,000-43,605,000) and fluid intelligence (chr15:43,645,000-43,650,000), respectively.

### Mosaic deletions associated with leukemia

One advantage of read depth is its ability to detect somatic CN changes that are present in a fraction of cells. We identified several loci showing apparently mosaic deletions associated with strongly increased risk (>100-fold) of two types of leukemia (Supplementary Figure 10). These included four loci associated with risk of chronic lymphocytic leukemia (CLL): the *IGKV* gene cluster at 2p11.2 (p = 5.9 x 10^-493^), the *IGHV* gene cluster at 14q32.33 (p = 1.5 x 10^-545^), the *IGLV* gene cluster at 22q11.22 (p = 3.0 x 10^−269^) and a >20Mb region at 13q14.11-q21.2 (p = 2.2 x 10^-317^). In addition, we identified known mosaic deletions of an ∼260-kb region encompassing *TET2* associated with an ∼145-fold increased risk of chronic myeloproliferative disease (p = 1.4 x 10^-10^)^31^. Of note, coding point mutations of *IGLL5*, which lies within the *IGLV* locus, have previously been identified as one of the most frequent coding mutations in CLL^32^. Consistent with this, the strongest association signal for CLL risk we observed at the *IGLV* locus occurred for a region overlapping *IGLL5*. Thus, our observations extend the spectrum of *IGLL5* mutations causing CLL to include genomic deletions of 22q11.22.

### Non-coding CNVs

PheWAS identified several intergenic CNVs that showed strong effects on traits known to be regulated by nearby genes, suggesting that these CNVs act via dysregulation of cis-linked expression. For example, coding mutations in *MC4R* are the most common cause of monogenic obesity^33,34^. We identified a rare non-coding deletion (MAF ∼0.0002) located ∼100kb upstream of *MC4R*, carriers of which were an average of ∼14 kg heavier than controls (p = 9.5 x 10^-11^) (Supplementary Figure 11).

Other similar examples include an ∼20-kb deletion (MAF ∼0.018) located ∼100kb upstream of *TACR3* that was associated with altered secondary sexual characteristics. Mutations in *TACR3* cause hypogonadotropic hypogonadism^35^ and female carriers of the deletion showed age of menarche delayed by an average of ∼4 months compared to controls (p = 7.1 x 10^-39^), while in male carriers, testosterone levels were reduced by an average of ∼5% (p = 4.8 x 10^-15^) (Supplementary Figure 12). Similarly, a deletion located ∼80-kb upstream of *BMP5*, a gene with known roles in bone morphogenesis^36^, was associated with increased heel bone mineral density (p = 1.1 x 10^-27^) (Figure 6).

**Figure 6.**
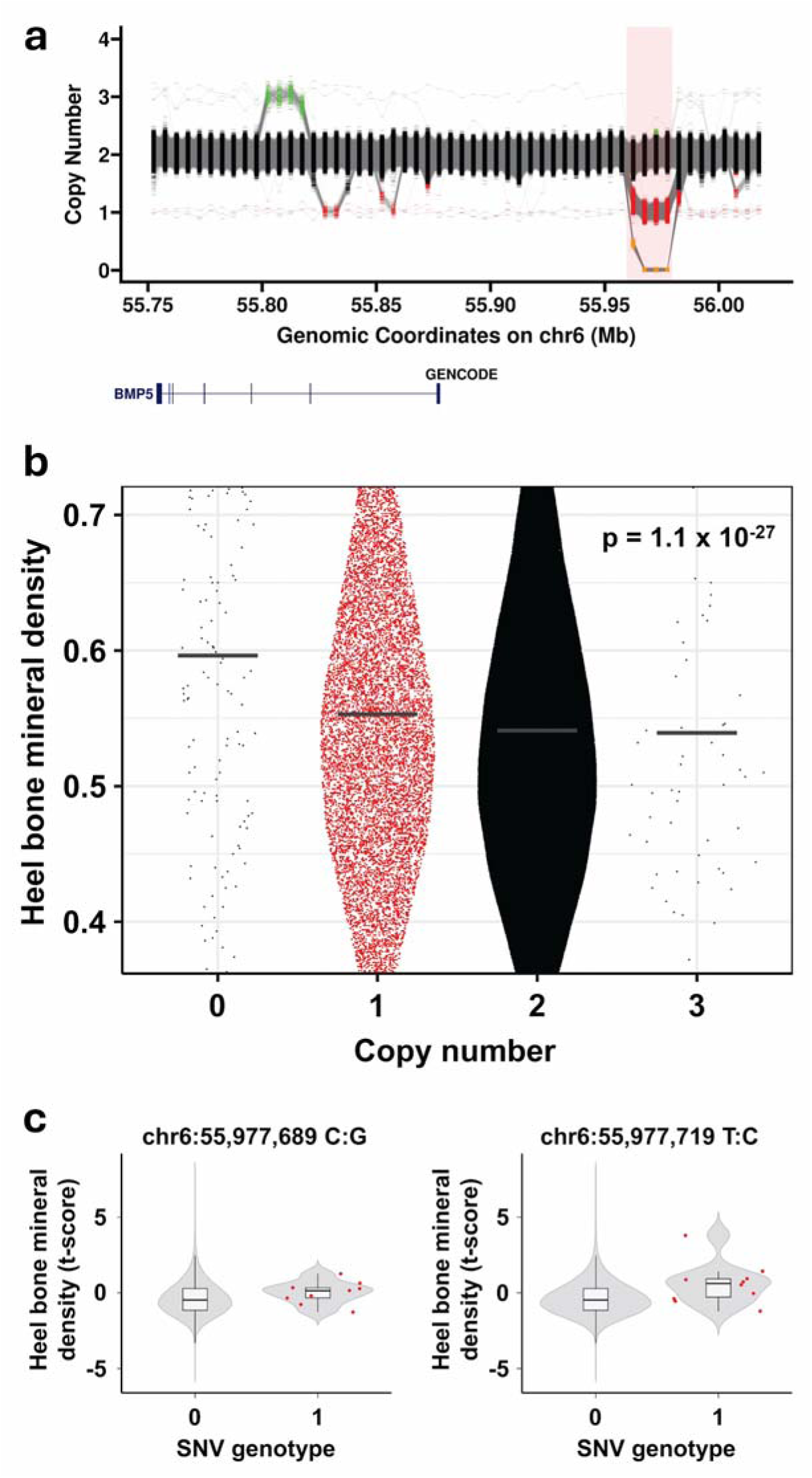
Disruption of non-coding regulatory elements by either CNVs or rare constrained SNVs yields similar trait associations. (**a** and **b**) PheWAS identified a non-coding deletion ∼80-kb upstream of bone morphogenetic protein 5 (*BMP5)* associated with increased heel bone mineral density (p = 1.1 x 10^-27^), a region including several annotated cis-regulatory elements. **(c)** Using burden tests, we identified a cluster of rare constrained SNVs within the deletion region that overlapped one of these regulatory elements and was associated with increased heel bone mineral density t-score (p = 7.1 x 10^-5^). In **(a)**, the red shaded region indicates the location of the deletion. In **(b)**, horizontal black bars indicate the mean of each distribution, with data shown for the 5-kb bin with the strongest association (chr6:55,970,000-55,975,000). In **(c)**, we show data for two representative SNVs. Within each violin, box limits indicate the 25^th^ and 75^th^ percentiles while the internal black bars show the medians; whiskers extend 1.5 times the interquartile range from the 25^th^ and 75^th^ percentiles. Red points show individual values where n < 50.

In some cases, these non-coding CNVs encompass annotated regulatory elements of an adjacent candidate gene for the associated trait, suggesting a likely mode of action for these CNVs. For example, *CXCR4* is a chemokine receptor, mutations in which cause an immunologic disorder characterized by neutropenia^37^. We detected a rare duplication (MAF ∼0.00005) located ∼110-kb upstream of *CXCR4* that overlaps an annotated enhancer of *CXCR4*^38^ and associates with altered monocyte count (p = 1.9 x 10^-10^). Similarly, mutations in *IRF8* cause autosomal dominant Immunodeficiency 32A^39^. We identified a rare deletion (MAF ∼0.001) located ∼70kb downstream of *IRF8* that includes an annotated enhancer of *IRF8*^38^ and was associated with significantly altered monocyte count (p = 2.8 x 10^-14^) (Supplementary Figure 13).

### Rare constrained SNVs within non-coding CNV regions show similar trait associations

Based on the observation that some non-coding CNVs likely exert trait effects via disruption of cis-regulatory elements, we hypothesized that single nucleotide mutations within the same loci might also have similar phenotypic effects. Using metrics of genomic constraint^40,41^, we defined potentially functional SNVs within six candidate non-coding CNVs identified via PheWAS and, using burden tests, searched for clusters of rare constrained SNVs that associated with similar traits to those seen in CNV carriers. We identified significant (Bonferroni-corrected p<0.05) associations between clusters of rare SNVs within an enhancer of *CXCR4* that associated with monocyte count (p = 1.2 x 10^-5^) and within a cis-regulatory element upstream of *BMP5* that associated with heel bone mineral density t-score (p = 7.1 x 10^-5^) (Figure 6c, Supplementary Figure 14, Supplementary Table 3). These results indicate that, at some loci, disruption of regulatory elements, either by large deletions or through point mutation of constrained nucleotides within the same locus, can cause similar phenotypic effects.

### PheWAS in non-European ancestries

The UKB includes a small proportion of individuals of non-European ancestry. We performed additional PheWAS using individuals of African (AFR, n=6,165) and South Asian (SAS, n=3,628) ancestry, respectively, representing the two largest non-European ancestry groups in UKB. Despite the small sample sizes limiting statistical power, these analyses identified 28 significant (Bonferroni p < 0.05) CNV:trait pairs in AFR and 15 in SAS, including several signals that were not identified in individuals of EUR ancestry (Supplementary Figures 15 and 16, Supplementary Tables 4-7). AFR-specific signals included multiple CNVs at the *HBB* locus associated with sickle cell disease and erythrocyte distribution width, CNVs of *F8* associated with hemoglobin A1c levels and red blood cell traits, and deletions overlapping *LPAL2* associated with increased lipoprotein A levels.

## Discussion

In the past two decades, > 45,000 SNV-based GWAS have been performed, providing deep insights into the role of SNVs on human traits^42^. In contrast, progress in defining the contribution of CNVs genome-wide to human phenotypic variation has been relatively limited. By harnessing read depth as a simple but effective metric to genotype CNVs and assessing their relationship to > 13,000 traits, we identified diverse trait associations with hundreds of CNVs. Notably, we analyzed a much larger number of phenotypes than have been used in most prior studies^5–7^, a factor which contributed to the scale of our findings. We predict that extension of this framework to additional biobank datasets with available GS data will lead to many additional discoveries.

The use of read depth as a genotyping tool has limitations, such as low sensitivity for detecting sub-kilobase CNVs, an inability to detect balanced rearrangements and a lack of allelic and breakpoint information. Contrastingly, read depth from high (≥30x) coverage GS is highly accurate for genotyping CNVs ≥ 5-kb in size and notably allows genotyping for all types of CNVs and across the entire allele frequency spectrum, including complex events such as large multiallelic repeats and mosaic CNVs. This contrasts with many other short-read and array-based approaches that tend to perform poorly in complex genomic regions^43^ and imputation-based methods that typically have low accuracy for typing rare or recurrent events^44^. Another advantage of read depth is its speed and low cost, making it attractive for use in large cohorts. For example, we were able to generate genome-wide CN estimates for the entire UKB cohort (∼490,000 genomes) for a total cloud compute cost of only ∼$4,000 (< $0.01 per individual). In contrast, running pipelines such as *GATK-SV* typically costs ∼$2-2.5 per individual^45^. Finally, a major advantage of our method for use in association studies is that, by summarizing CN estimates per genomic region, our methodology effectively integrates data from multiple different sizes and types of CNV that overlap a locus, outputting a simple metric of total CN per interval that can be easily utilized in downstream analyses. In contrast, conventional genotype formats for structural variants often result in considerable complexity at some loci, with multiple different alleles, breakpoints and SV types annotated within a region that can confound their effective use in association studies.

Although we utilized three different association models (additive, dosage sensitive and recessive) the majority of signals we detected (76%) showed the strongest association using an additive model, indicating that most CNV effects can be detected using standard linear association methods. Notably, we detected several associations with recessive disease genes (*e.g.* homozygous deletions of *STRC* with hearing loss and deletion of X-linked opsin genes associated with eye traits in males) and dosage sensitive loci (*e.g.* large CNVs at 1q21.1, 15q13.3 and 22q11) that showed the strongest signals when using an association model consistent with their known inheritance pattern, indicating that our use of alternative disease models yielded increased power at some loci.

Our study provides a deep sampling of CNVs in the human population, representing a valuable resource to the community for many types of genomic study. While access to individual genotypes are restricted to approved UKB users, we have made our complete dataset of CN frequencies per 5-kb window together with significant trait associations derived from PheWAS available as public tracks in the UCSC Genome Browser (see Data Availability).

## Acknowledgements

This research has been conducted using the UK Biobank Resource under Application Number 82094. This work was supported by NIH grant AG075051 to A.J.S. and through the computational resources and staff expertise provided by Scientific Computing at the Icahn School of Medicine at Mount Sinai, supported by the Clinical and Translational Science Awards (CTSA) grant UL1TR004419 from the National Center for Advancing Translational Sciences and by the Office of Research Infrastructure of the National Institutes of Health under award number S10OD026880. The content is solely the responsibility of the authors and does not necessarily represent the official views of the National Institutes of Health. The funders had no role in study design, data collection and analysis, decision to publish or preparation of the manuscript. We thank Don Conrad for useful suggestions during the development of this work.

## Author Contributions

P.G. designed bioinformatics pipelines and performed data analyses. A.M.T., B.J. and M.S. performed data analyses. A.J.S. conceived the study, performed data analyses, supervised the project and drafted the manuscript. All authors reviewed and approved the final draft.

## Methods

### Ethics statement

Research performed in this study complies with all relevant ethical guidelines and informed consent for genetic research was obtained from all participants. Collection of UKB data was approved by the Research Ethics Committee of the UKB and protocols for UKB are overseen by The UKB Ethics Advisory Committee, see https://www.ukbiobank.ac.uk/ethics/.

### UK Biobank

UKB data^46^ were obtained under application 32568. From the set of 490,416 individuals with Illumina 150-bp paired-end GS data, we first removed samples that we were notified had withdrawn their consent for inclusion in the study or, using GS quality metrics, had values ≤ 2 for either “NRD genotyping” (UKB data field f32063) or “Freemix verify BAM ID” (f32062). We also utilized pairwise kinship coefficients provided by the UKB and, for samples with 2^nd^ degree relationships or higher (kinship coefficient > 0.0883), we retained a single unrelated individual.

### Ancestry prediction

We performed prediction of genetic ancestry in UKB individuals using available genotype data derived from SNV arrays based on a set of linkage disequilibrium (LD)-pruned, QC-filtered SNVs. Bi-allelic SNVs were filtered using *plink* (v1.9)^47^ to retain those with minor allele frequency (MAF) > 0.05, genotyping rate > 99% and Hardy-Weinberg equilibrium (HWE) p > 0.0001, before LD pruning with parameters --indep 50 5 1.5. We performed principal component analysis (PCA) on the set of LD-pruned SNVs from the UKB using *GCTA* (1.93.2b)^48^, together with a matching set of variants from the 1000 Genomes cohort^49^ as a reference panel. The first 10 principal components (PCs) were generated for the 1000 Genome dataset and UKB data were projected onto these PCA loadings. We then applied a random forest classifier to assign each UKB individual to one of five major ancestry groups: EUR, AFR, AMR, SAS and EAS. Where the assignment probability was < 0.5, the individual was labeled "undefined" and removed from further analysis (Supplementary Figure 17).

To assess fine-scale ancestry within each predicted major ancestral group, we performed within-group PCA using *GCTA* without an external reference panel. As EUR individuals comprised ∼94.3% of the UKB cohort, we randomly divided unrelated samples into three sets to reduce computational burden. PCs for one EUR set were calculated using *GCTA* and SNV data from the other two EUR sets were projected onto these PCs. The centroid for the ancestry group was calculated and outlier samples, identified by distance from the centroid, were excluded^50^. Samples with mismatches between self-reported ancestry and predicted ancestry, as well as those with low assignment probability or multiple self-reported ancestries across visits, were excluded from further analysis. We also removed individuals of AMR and EAS ancestry due to small sample sizes.

### Calculation of copy number estimates

CN estimates were generated from normalized read depth (RD) of Illumina genome sequencing (GS) data. The genome (hg38) was divided into consecutive non-overlapping 1-kb bins and the read depth for each 1-kb bin was calculated using *mosdepth* (v0.3.3)^51^ on the UKB DNAnexus Research Analysis Platform. To correct for potential bias due to variation in local GC-content (Supplementary Figure 18), a correction factor based on the approach of Yoon *et al.* was applied^52^. Specifically, a GC correction factor was derived for each 1-kb bin, as follows:

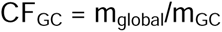

where m_GC_ is the median read count for all bins with a specific GC content (grouped in 1% intervals, *e.g.*, 0– 1%, 1–2%) and m_global_ is the global median read depth for all 1-kb bins. For GC content bins with fewer than 100 entries, adjacent bins were used to calculate the median, while, for bins with ≤10% GC, we used the median of all 1-kb bins with GC content ≤10%. We utilized the median to reduce the influence of extreme values.

Subsequently, raw read depths were normalized and converted into relative CN estimates as follows:

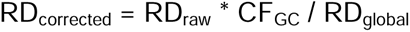

where RD_raw_ is the raw read depth for a bin and RD_global_ is the global mean RD across all bins. RD_corrected_ represents the relative CN for a 1-kb bin with respect to the reference genome. This adjustment corrected for GC bias and harmonized read depth values across the genome. Given that the accuracy of CN estimates based on read depth are related to the size of the region being assayed, for downstream analysis, we summarized data into non-overlapping 5-kb bins. We utilized 5-kb bins as we found that error rates increased significantly for smaller bin sizes, while the use of larger bins yields reduced resolution.

### Correcting for genomic waves and batch effects

Genomic waves are systematic biases that can occur in CN estimates from both array and sequencing data, impacting accurate CN quantification^7,53^. To reduce the effect of genomic waves, we divided the genome into consecutive non-overlapping 2-Mb windows and calculated a genomic wave correction factor for each 2-Mb window per sample based on the constituent 5-kb bins that met the following criteria:

1. The mean CN across all individuals was between 1.8 and 2.2.
2. The difference between the 0.01^st^ and 99.99^th^ percentiles was < 0.9.
3. Sample CN values fell within the 0.01^st^ and 99.99^th^ percentiles.

In addition to the above requirements, to ensure that we used for this normalization only those 5-kb bins with robust and stable CN estimates, we excluded any 5-kb bins that overlapped with segmental duplications (“Repeats > Segmental Dups” track in UCSC Browser), centromeres (“Mapping and sequencing > Centromeres” track in UCSC Browser), low Complexity or satellite sequences (sub-tracks with corresponding name in “Repeats > Repeat Masker” track in UCSC Browser), or simple repeats (“Repeats > Simple Repeats” track in UCSC Browser). The genomic wave correction factor was calculated per 2-Mb window as the mean difference of each individual CN from 2 and was used to adjust CN for all 5-kb bins (including those excluded above) within that 2-Mb window. In cases where, after filtering, there were < 50 5-kb bins remaining within a given 2-Mb window (typically regions adjacent to centromeres or telomeres), here we utilized the correction factor from the adjacent 2-Mb window(s) to calculate the genomic wave correction factor (Supplementary Figure 19). The correction factor was then applied on a per-sample basis to adjust CN estimates for all 5-kb bins within the corresponding 2-Mb window.

At some loci, we observed batch effects in CN distributions associated with both sequencing center (deCODE or Sanger Center) and phased releases of the UKB GS data (Vanguard, initial 200,000 GS data release, final 300,000 GS data release) (Supplementary Figure 20). To adjust for these effects, for each 5-kb window, median CN was calculated for each batch and adjusted to the global median CN of all samples:

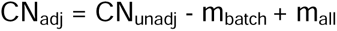

where CN_unadj_ is the CN normalized after correcting for genomic waves, m_btach_ is the median CN of individuals within the sequencing batch to which the sample belongs to and m_all_ is the median CN across all individuals and batches. This median adjustment reduced inter-batch variability by normalizing each batch to a global median while preserving underlying CN distributions within each batch.

### Outlier removal based on constrained genes

We implemented three additional QC steps to remove technical outliers. For this, we selected 200 highly constrained autosomal genes (pLI scores > 0.9)^54^ that also showed very low CN variance in samples from the Human Genome Diversity Panel^55,56^. Considering all 5-kb bins overlapping the coding regions of these genes, we calculated the mean CN per gene per individual. In the first step, individuals that were consistent outliers, defined as those that showed CN estimates ≥ 4 standard deviations from the mean for ≥ 10 of these 200 highly constrained genes were removed from further analysis (Supplementary Figure 20). Second, after dividing individuals based on the sequencing center and data release batch, we performed PCA using estimated CN of these 200 constrained genes as input and removed outliers based on the top six PCs (Supplementary Figure 21). Finally, we removed individuals with extreme outlier GS insert size, calculated per sequencing center and batch. These quality filtering steps resulted in significant improvements to downstream CN clustering and, therefore, improved genotype quality (Supplementary Figure 22).

### Identification of autosomal and sex chromosome aneuploidy

As the presence of either constitutional or mosaic aneuploidy confounds read-based CN estimates, we set out to identify and remove individuals with evidence of aneuploidy using read depth estimates per chromosome. To increase accuracy for aneuploidy detection, for this analysis we excluded all 1-kb bins overlapping regions of Segmental Duplication (“Repeats > Segmental Dups” track in UCSC Browser), Simple Repeats (“Repeats > Simple Repeats” track in UCSC Browser) and Low Complexity regions (sub-track “Low complexity” in “Repeats

> Repeat Masker” track in UCSC Browser). For chromosomes X and Y, bins overlapping the pseudoautosomal regions (PARs) were also excluded.

We performed prediction of sex chromosome complement using the relative ratios of chromosome X:autosomes and chromosome Y:autosomes based on median read depth per sample per chromosome. We excluded individuals with a mismatch between self-reported gender and predicted sex, as well as those with predicted sex chromosome aneuploidy (*e.g.*, 45,X, 47,XXX, 47,XXY.) (Supplementary Figure 23).

Aneuploidy for each autosome was assessed by computing the ratio of median read depth per chromosome to the median read depth across all autosomes in each individual. Individuals showing noticeable deviations from the chromosome-specific population mean ratios, indicating either mosaic or constitutional aneuploidy, were manually flagged and removed from further analysis. (Supplementary Figure 24). After implementing all quality filters, we retained CN estimates for 405,362 quality-filtered unrelated individuals of European ancestry that were utilized for PheWAS.

### Clustering into discrete copy numbers

The use of read depth yields an estimate of relative CN per within each individual compared to the hg38 reference genome. We chose to summarize data in the format of 5-kb bins tiled across the genome, as this gave a good balance between accuracy and resolution, although other bin sizes could be used. Smaller bins yield improved resolution for typing smaller CNVs but with increased noise. Conversely, the use of larger bin sizes reduces noise but at the expense of decreased resolution. CN estimates per bin are continuous variables that include error due to the inherent stochasticity of shotgun GS, batch effects and other potential biases in the data. In order to convert these relative CN estimates to discrete CNs that more accurately reflect the nature of the genome (0, 1, 2, 3, *etc.*), for each 5-kb bin, based on the observed data, we determined the optimal clustering model to best represent the potential variability in reference CNs. For instance, for unique regions of the genome, a reference copy of one would yield clustering at approximately integer values (0, 1, 2, 3, *etc.*). In contrast, for a region of hg38 that is present in the genome in two copies, the data would generally better fit with a CN model of two, with clusters at increments of ∼0.5 (0, 0.5, 1, 1.5, 2, *etc.*). We tested models up to a maximum of 10 reference copies, which typically yielded clusters spaced by ∼0.1 (0, 0.1, 0.2, 0.3, *etc.*), reflecting highly multiallelic loci that are often present at high CN.

For each CN model, the centroid for each cluster was initialized based on the model selected. For example, the initial centroids for a model with reference copy of 1 were set to 0, 1, 2, 3 and so on. Cluster centroids were then refined using the observed CN distribution for each 5-kb bin. For clusters with fewer than 50 data points, the mean of the points was used as the centroid, whereas for clusters with 50 or more data points, the density peak was chosen as the centroid. Clusters with fewer than 20 data points that showed low separation from other clusters, defined as those where their difference between centroids was less than half of model’s cluster spacing, were merged with their nearest neighboring cluster to avoid over-fragmentation. Following the refinement of centroids, data points were re-clustered based on the updated centroids.

For each different CN model tested, we evaluated the clustering quality and selected the optimal CN model using Silhouette scores (v0.0.1)^57^. Silhouette scores provide values ranging from -1 (misclassification) to +1 (perfectly clustered). In cases where the Silhouette score for a cluster comprising a single data point was negative, then that data point was assigned to the nearest neighboring cluster. After reassignment, the sum of individual Silhouette scores provided an overall score for the model. Where only a single individual was present in a cluster (*i.e.*, a very rare CN state), a pseudo data point was added to enable score calculation. The clustering model that yielded the highest Silhouette score was selected as the optimal model, with the total number of clusters used as a reference for defining CN group structure. Finally, a hierarchical tree was generated to represent the clustering structure and the previously determined optimal number of clusters was used to cut the tree, thereby defining final CN clusters, resulting in discrete CN calls per individual for each 5-kb bin. These discrete CN calls were utilized for PheWAS.

Overall, our CNV profiling pipeline proved very efficient, with compute costs for running *mosdepth*, performing GC-correction, wave correction and clustering costing ∼$0.008 - 0.01 per individual. This enabled us to generate genome-wide CN estimates for the entire UKB cohort for a total cost of ∼$4,000.

### Steps for clustering

#### 1. Model Initialization

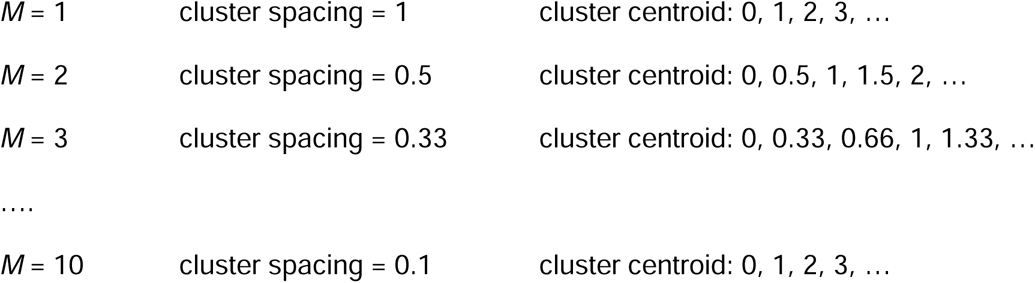

where *M* implies reference copy for the model

#### 2. Refine Cluster Centroids

For each centroid, *c* ∈ *C_M_*

- If cluster has < 50 data points, update *c* → mean of assigned points
- If cluster has ≥ 50 data points, update *c* → density peak

#### 3. Merge clusters **if:**

Cluster has < 20 data points AND

Distance between centroids < 0.5 x model’s cluster spacing.

#### 4. Re-clustering

Re-assign data points to the nearest refined centroids.

#### 5. Evaluate clustering quality

- Compute Silhouette score *S*_/, M_ ∈ *[-1, 1]* for each sample *i* and model *M*.
- If *S*_/, M_ < 0, reassign the sample to the nearest cluster.
- If cluster size = 1, add pseudo point to compute *S*_/, M_
- Compute total model score *S*_M_ *=* ∑*S*_/, M_

#### 6. Select optimal model

*M* = max S_M_*

#### 7. Final Clustering

- Perform hierarchical clustering, cut tree at |C_M*_| clusters
- Use discrete clusters for PheWAS

### Estimating accuracy of copy number calls using twin pairs

We identified 146 pairs of presumptive MZ twins of European ancestry in the UKB cohort based on pairs of individuals with kinship coefficients ≥ 0.35 and with concordant age and sex. Within each pair, we used data from 5-kb bins located on chromosomes 1-22 to assess concordance of genotypes after clustering into discrete CNs. For each 5-kb bin, we calculated how many times the twin pairs were clustered in the same CN group (*i.e*., concordant genotypes) or different CN groups (*i.e.*, discordant genotypes).

### Filters to remove unreliable genomic loci

We observed that CN estimates from read depth were often highly unstable when certain repetitive elements or genome assembly errors were present within a locus. Therefore, we removed from analysis any 5-kb bin that had ≥ 500 bp of overlap with any of the following annotations: L1P and/or L1HS elements (based on the LINE sub-track in “Repeats > Repeat Masker” track in UCSC Browser), satellite elements (based on the Satellite sub-track in “Repeats > Repeat Masker” track in UCSC Browser), rRNA elements (based on the RNA repeats sub-track in “Repeats > Repeat Masker” track in UCSC Browser), AT-rich tandem repeats (defined as those with motifs comprised of ≥90% AT content based on the “Repeats > Simple Repeats” track in UCSC Browser), genome assembly gaps (based on the “Mapping and sequencing > Gap locations” track in UCSC Browser) and centromeric-like pentamer repeats with the motifs TTCCA, TCCAT, CCATT, CATTC or ATTCC, or their reverse complements (based on the “Repeats > Simple Repeats” track in UCSC Browser). Finally, we excluded any 5kb bin that showed either a Silhouette score < 0.5 or had extremely high CN, defined as those with maximum CN > 120.

### PheWAS analysis in the UKB

We utilized phenotype data for individuals in the UKB derived from phenotype fields containing ICD10 codes and quantitative, categorical and binary traits, accessed through UKB application number 82094. Before performing association testing, phenotype data were processed as follows:

1. For phenotypes with multiple categories, *e.g.*, ICD10 codes, we considered each category as a separate binary trait. If a subject had multiple entries for the same category, they were assigned as “1” (*i.e.*, positive for that trait) if any of the instances were positive. We removed from analysis any categories labeled as “Prefer not to answer”, “Unsure” or “Do not know”.
2. In addition to using each individual ICD code as a separate trait, ICD codes were also grouped into higher level codes to yield additional summary traits with increased sample size. For example, the ICD10 code for schizophrenia is F20, which is composed of 10 different sub-codes (e.g. F20.0 “Paranoid schizophrenia:, F20.2 “Catatonic schizophrenia”, F20.9 “Schizophrenia, unspecified”, etc), with each subcategory containing between 4 and 894 individuals. Here, we created a new summary code for “Schizophrenia_group” comprising all sub-categories under F20 which included 1,485 individuals who were positive for any subtype of schizophrenia. Similar groupings were performed for any higher-level ICD code that comprised multiple subtypes.
3. For phenotypes with > 2 categorical outcomes, each outcome was assigned an integer value and these were analyzed as quantitative traits. *e.g.*, the phenotype “Alcohol intake frequency” included six categories: "Daily or almost daily", "Three or four times a week", "Once or twice a week", "One to three times a month", “Special occasions only” and “Never”. Here, each sample was assigned an integer value ranging from 1 to 6, with each value corresponding to the six ordered frequencies for this phenotype. Categorical traits with just two separate categories were considered as binary traits.
4. For quantitative traits with integer values, where a subject had multiple instances, we utilized the mean of all instances rounded to nearest integer
5. For quantitative traits with continuous values, where a subject had multiple instances, we utilized the mean of all instances.

For all quantitative traits, we applied a rank based inverse normal transformation and required a minimum sample size of 5,000 phenotyped individuals and standard deviation of the trait ≥ 0.001 to be utilized for PheWAS. For binary traits, we required a minimum sample size of at least 100 individuals with and 100 individuals without the trait to be utilized for PheWAS. After these filters, we utilized data from a total of 13,215 traits for PheWAS (Supplementary Table 8).

Association analysis was performed using *REGENIE* (v3.4.1)^58^, incorporating covariates of sex, age, GS insert size and the top three PCs derived from analysis of SNVs to account for ancestry. For binary traits, we utilized the Firth approximation function to reduce the type I error rate.

For PheWAS of chromosome X, to ensure accurate genotyping of CNVs, we applied more stringent filtering to remove samples with any evidence of possible mosaic aneuploidy of chromosome X, which is commonly observed in blood cells and tends to increase with age^59^. Based on the calculated relative ratio of chromosome X:autosomes (detailed above), we removed additional individuals that showed outlier values, resulting in a final set of 218,308 females and 175,133 males used for PheWAS of chromosome X. Males and females were then analyzed separately. Instead of using sex as a covariate, we incorporated the ratio of mean read depth on chromosome X (in females) or on chromosome Y (in males) to the autosomes as a covariate to account for any residual mosaic aneuploidy of the sex chromosomes. P-values for males and females were combined using a weighted Stouffer’s Z-score method, where the number of samples in each group were used as weights.

### Additive, recessive and dosage sensitive models

We required that 5-kb bins showed StDev of raw (unclustered) CN estimates ≥ 0.5, that the difference between the maximum observed CN and minimum raw CN was ≥ 0.9 and that there were ≥ 20 individuals defined as having a non-modal CN.

For the additive model, where CN is assumed to be linearly associated with the trait, we first computed the mean CN for each cluster within each 5-kb bin. These values were then rescaled to a 0-2 range to represent dosage values. The data were formatted into VCF files, with dosages included as the DS field in the genotype data. Finally, we used *plink2* (v3 May 22)^60^ to convert the VCFs into PGEN format for input into *REGENIE*.

In addition to the additive model, we also performed PheWAS using both recessive and dosage sensitive models to relate genomic CN to phenotype. These models were chosen as they reflect known biological mechanisms by which change in genomic CN can lead to a phenotype. For both the recessive and dosage sensitive models, we only considered those 5-kb windows with an underlying CN model of 1 or 2, while regions with CN models ≥ 3, indicative of an underlying multiallelic state, were excluded. As applied for the additive model, for both dosage sensitive and recessive models, we required that the difference between the maximum observed CN and minimum raw CN was ≥ 0.9 and that there were ≥ 20 individuals defined as having a non-modal CN.

Under the recessive model, we assumed that homozygous deletion of a region would associate with a trait, whereas all other CNs would not. Here, for each 5-kb window, individuals were considered to carry a homozygous deletion when they were assigned to the cluster with the lowest mean CN and where the mean CN value of that cluster was ≤ 0.5. Individuals in these clusters were assigned a genotype of “0/0”, while samples in all other CN clusters were assigned genotypes of “0/1”.

Under the dosage sensitive model, we assumed that either gain or loss of copies would associate with a trait, whereas the modal CN state would not. Here, for each 5-kb window, we identified the modal cluster containing the greatest number of individuals. Individuals in this modal cluster were assigned a genotype of “0/0” while samples with genotypes in all other CN clusters were assigned genotypes of “0/1”.

### PheWAS using non-European populations from the UKB

Using PCA of high quality autosomal SNVs, as described above, we identified 6,459 individuals of AFR and 3,825 individuals of SAS ancestry. CN profiling was performed as described above, generating new CN models for each 5-kb bin throughout the genome in both the AFR and SAS cohorts. After applying the same QC, filtering and data normalization methods as utilized for individuals of European ancestry, for PheWAS we utilized a set of high-quality CN calls for 6,165 and 3,628 unrelated individuals of AFR and SAS ancestry, respectively. PheWAS was performed using the same methods utilized for EUR samples, as stated above, except that the minimum sample size required to analyze quantitative traits was reduced to 1,000.

### Estimating a multiple testing correction

Given that many phenotypes in UKB are highly correlated, application of multiple testing corrections based purely on the number of traits tested tend to be overly stringent. Furthermore, as many CNVs are large, often spanning tens or hundreds of-kb, multiple contiguous 5-kb bins in the genome will often have highly correlated CN. Therefore, to estimate a better-calibrated Bonferroni threshold in our analysis, we calculated the number of nominally independent traits and CNV regions tested by calculating the pairwise Spearman correlation coefficient between all traits and between all 5-kb bins that were utilized in our PheWAS. Considering a threshold of R^2^ < 0.1 between any two traits or between any two 5-kb bins as indicative of independence, we calculated the minimal set of independent tests performed in our PheWAS. For CNV regions, we restricted correlation estimates to 5 kb bin pairs that were ≤ 10 Mb apart on the same chromosome. For traits, we used the cluster_louvain function in R that iteratively groups the traits and refined group membership based on the correlation coefficient. Based on this, in Europeans, we considered signals that passed an adjusted Bonferroni-corrected threshold of p < 3.01 x 10^-10^ (p=0.05 / 26,727 independent CNV regions / 6,215 independent traits) to indicate significant associations. In AFR and SAS cohorts, the reduced sample size meant that fewer CNVs and traits met the minimum counts necessary to be used in PheWAS, resulting in Bonferroni-corrected thresholds of p < 9.1 x 10^-9^ in AFR and p < 2.1 x 10^-8^ in SAS.

### Testing the influence of rare single nucleotide variants within non-coding CNV regions

For six non-coding CNV regions identified via PheWAS, we utilized burden tests^61^ to identify whether clusters of rare constrained SNVs were associated with the same or similar traits to those identified by CNV PheWAS. For each region, in the DNAnexus portal, we used *BCFtools (v1.21)*^62^ to extract SNVs from GS data for unrelated EUR UKB individuals without evidence of chromosomal aneuploidy. Multiallelic SNVs were converted to biallelic format and we retained those with MAF < 0.001, QUAL ≥ 30, GQ ≥ 20, DP ≥ 10 and call rate ≥ 0.9. Using *BEDTools (v2.31.0)*^63^, we annotated putatively constrained SNVs as those having either JARVIS^40^ scores > 0.49 or GERP^41^ scores > 1. Each non-coding CNV region was then divided into non-overlapping 500 bp windows and, for each window containing ≥ 5 SNVs, we applied six rare variant association methods (SKAT, SKAT-O, SKATO-ACAT, ACATV, ACATO, ACATO-FULL)^64^ and two joint tests (BURDEN-SBAT, BURDEN-ACAT), implemented in *REGENIE*, incorporating covariates of sex, age and the top five PCs derived from genome-wide SNVs. For sex-specific traits (*e.g.*, age of menarche), we performed analysis using samples of the relevant sex. Quantitative traits were subject to rank-based inverse normal transformation. We applied a Bonferroni correction based on testing 142 non-overlapping 500-bp windows.

### Statistics & Reproducibility

No statistical methods were used to pre-determine sample sizes, but our sample sizes were similar or greater to those reported in previous publications^65^. Data distributions were assumed to be normal but this was not formally tested. The experiments were not randomized. All data exclusions and characteristics of the populations used in this study are described in the Reporting Summary.

## Data Availability

UKB Data Showcase, https://biobank.ctsu.ox.ac.uk/crystal/search.cgi. CNV genotypes generated from the UK Biobank have been submitted as a Data Return to the UK Biobank, and will be made available in a future data release once processing of these is complete by the UK Biobank team.

CNV allele frequencies, copy number states and significant trait associations per 5-kb bin are available as tracks in the UCSC Genome Browser (https://genome.ucsc.edu/s/paras.mssm/UKBB_CN_tracks)

## Code Availability

Code utilized for this study is available as follows:

Global ancestry assignment, https://zenodo.org/doi/10.5281/zenodo.10820994

Copy number profiling using read depth, normalization and clustering, https://github.com/pgarg-tools/cn_estimator

Burden testing for non-coding variants, https://github.com/shadrinams/SNVBurdenTests

