## Supplementary Figures 1-24 for "A phenome-wide association study of CNVs genotyped from genome sequencing read depth in the UK Biobank"

**Supplementary Information**

**List of Supplementary Tables**

**Supplementary Table 1.** Results of PheWAS on chromosomes 1-22 in 405,362 Europeans.

**Supplementary Table 2.** Results of PheWAS on chromosome X in 405,362 Europeans.

**Supplementary Table 3.** Results of testing rare constrained SNVs within six non-coding CNV regions identified via PheWAS

**Supplementary Table 4.** Results of PheWAS on chromosomes 1-22 in 6,165 Africans.

**Supplementary Table 5.** Results of PheWAS on chromosome X in 6,165 Africans.

**Supplementary Table 6.** Results of PheWAS on chromosomes 1-22 in 3,628 South Asians.

**Supplementary Table 7.** Results of PheWAS on chromosome X in 3,628 South Asians.

**Supplementary Table 8.** All phenotypes used for PheWAS, including counts of individuals with data for each trait per ancestry.

**Supplementary Figures**

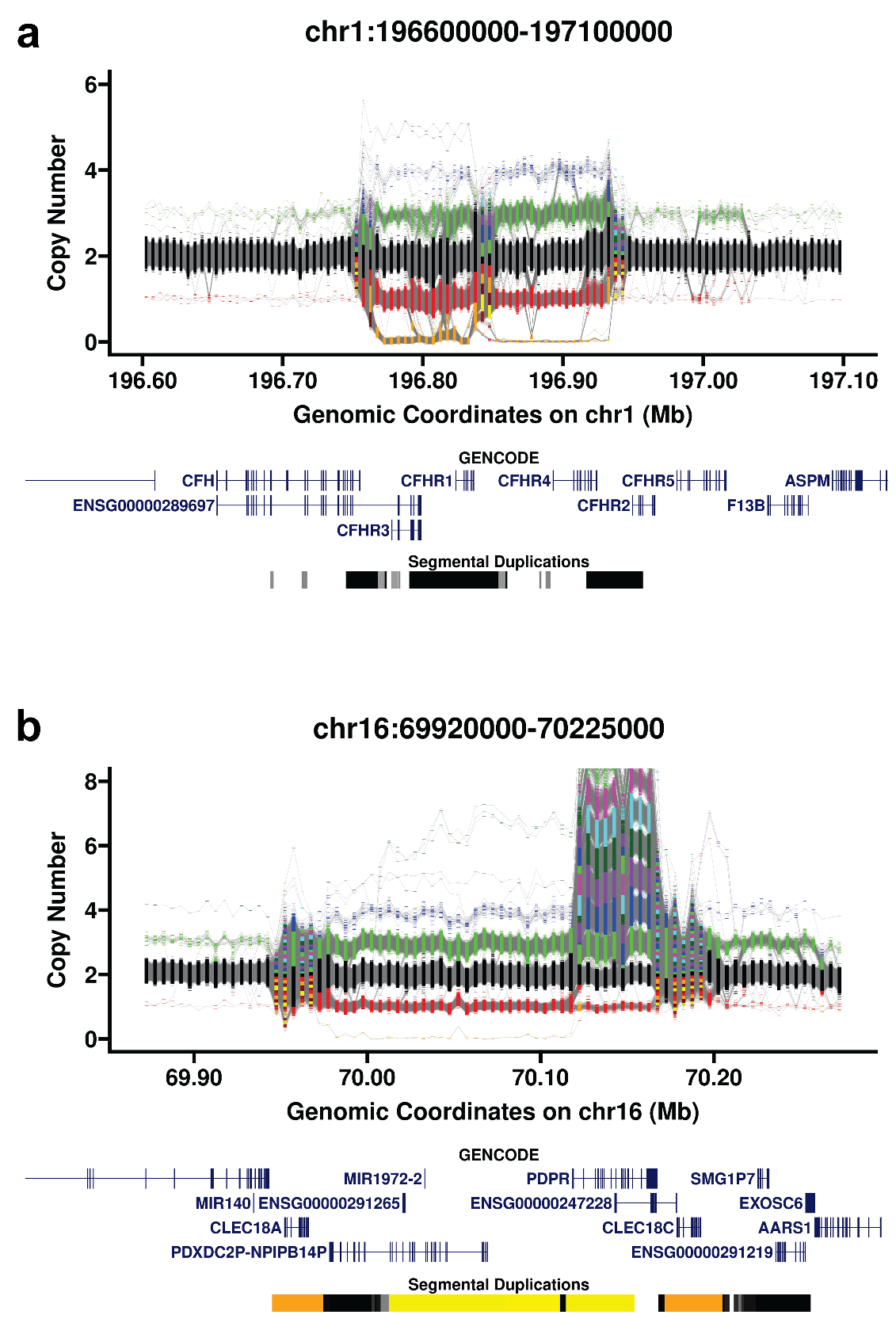

**Supplementary Figure 1. Example CN calls at two genomic loci showing high variability.** (a) The Complement Factor H locus and (b) The pyruvate dehydrogenase locus.

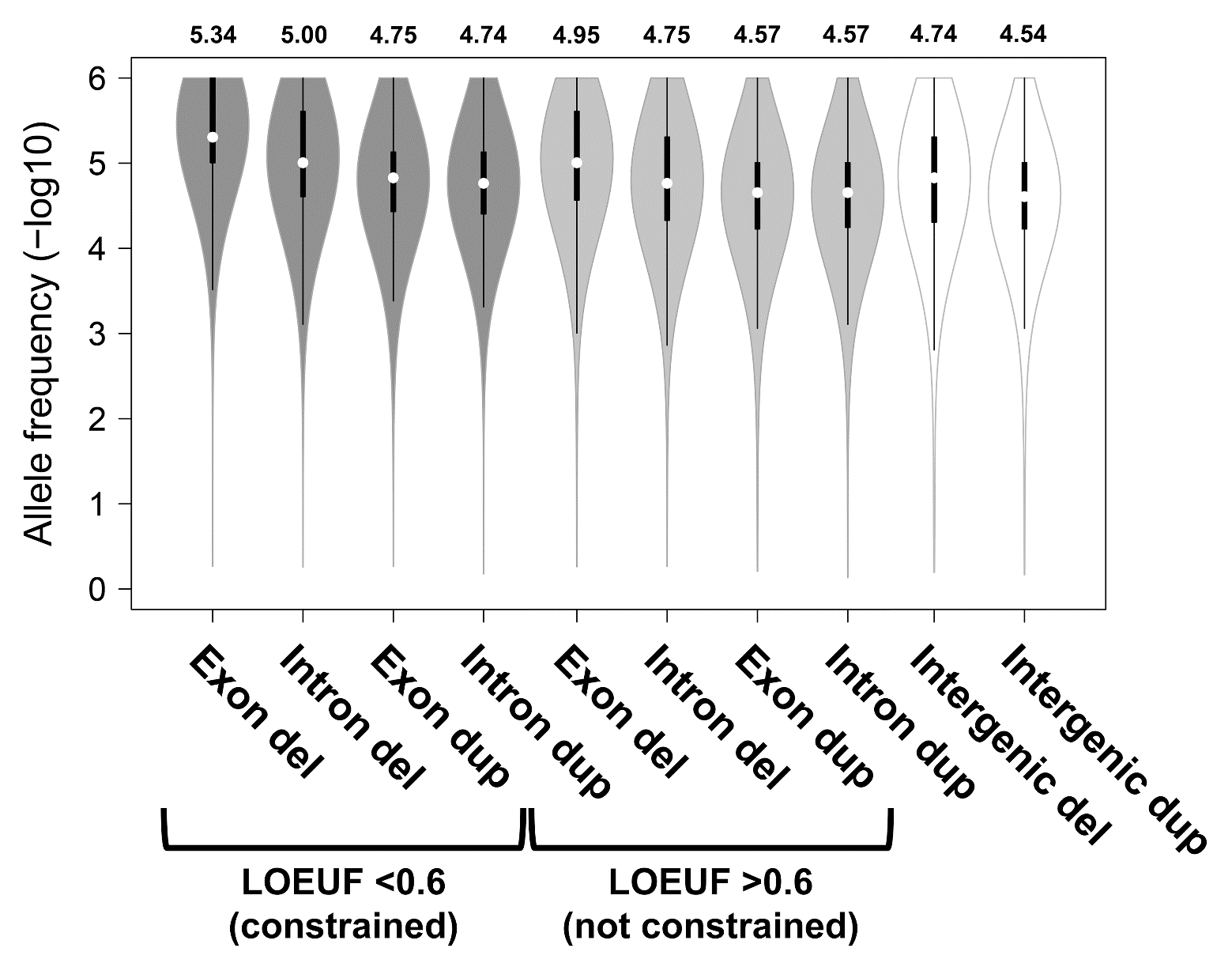

**Supplementary Figure 2. Allele frequency of CNVs varies by type, genic location and gene intolerance to loss-of-function variation.** Plot shows allele frequency by CNV type and location, calculated using all autosomal 5kb bins used for PheWAS that had copy number model = 1 (*i.e.* excluding multiallelic CNV loci), based on 405,362 unrelated, quality-filtered UKB participants of European ancestry**.** “Del” indicates deletions (copy number losses) while “dup” indicates duplications (copy number gains). RefSeq genes were annotated with LOEUF scores from gnomAD v4, with scores < 0.6 taken to indicate constrained genes^12^. “Exon” indicates 5kb bins that overlapped coding regions of a RefSeq gene, “intron” indicates all other 5kb bins that overlapped a RefSeq gene, while “intergenic” indicates 5kb bins that had no overlap with RefSeq genes. Within each violin, white circles show the medians, box limits indicate the 25^th^ and 75^th^ percentiles, whiskers extend 1.5 times the interquartile range from the 25^th^ and 75^th^ percentiles. Numbers above the plot show the mean of each violin.

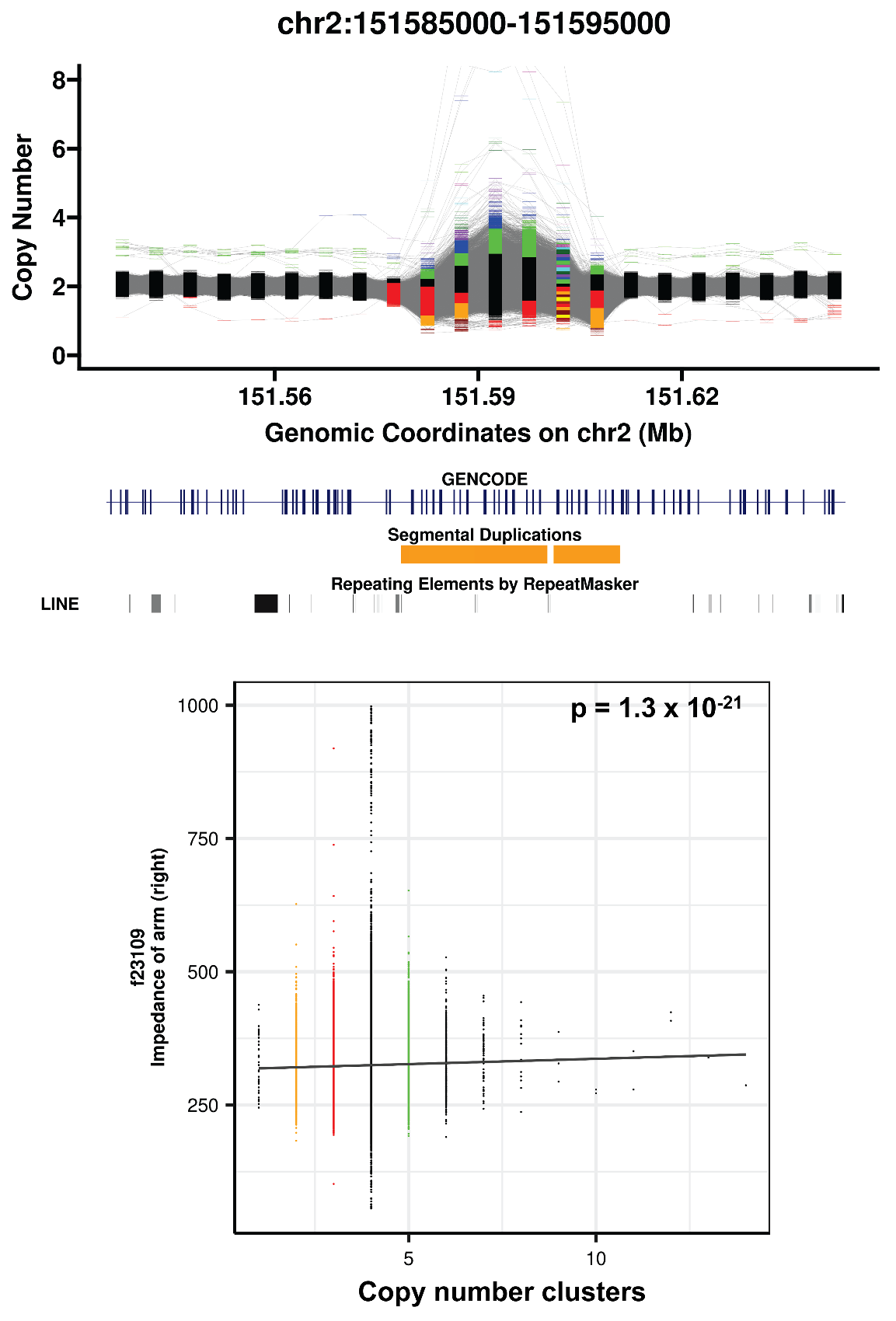

**Supplementary Figure 3. Copy number of a region in *NEB* is associated with arm impedance, a measure of muscle mass from bioelectrical impedance analysis.** Upper plot shows CN estimates per 5 kb bin across the *NEB* locus within 2q23.3. Lower plot shows impedance of right arm (y-axis) versus copy number state for the 5 kb bin with the strongest association (chr2:151,585,000-151,590,000). Solid black line shows the regression slope.

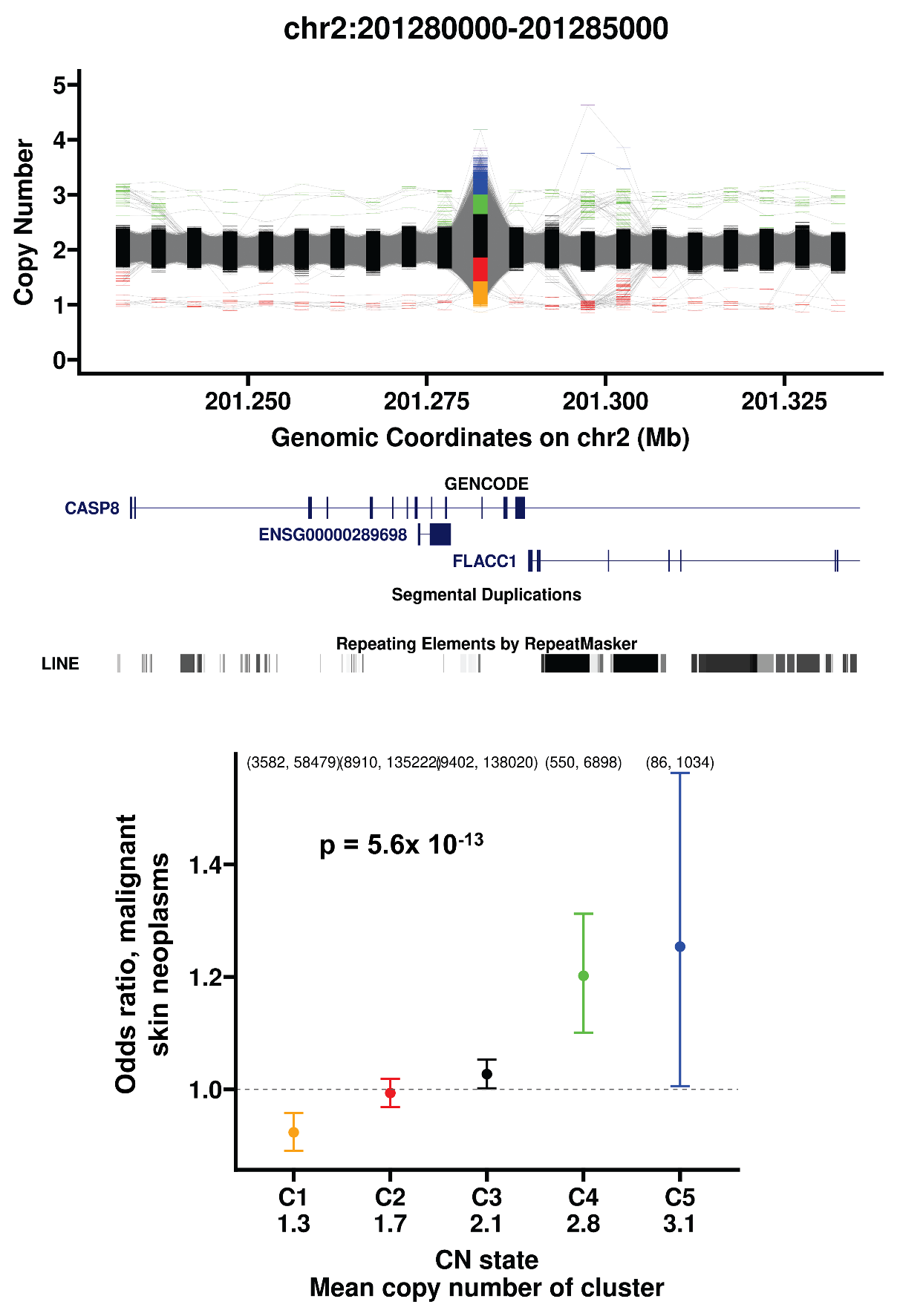

**Supplementary Figure 4. Copy number of an intronic region in *CASP8* is associated with risk of malignant skin neoplasms.** Upper plot shows CN estimates per 5 kb bin across the *CASP8* locus within 2q33.1. Lower plot shows odds ratio for malignant skin neoplasms (y-axis) versus copy number state for the 5 kb bin with the strongest association (chr2:201,280,000-201,285,000). Vertical bars represent 95% confidence intervals, with numbers in parentheses indicating the number of individuals per copy number state with and without the trait.

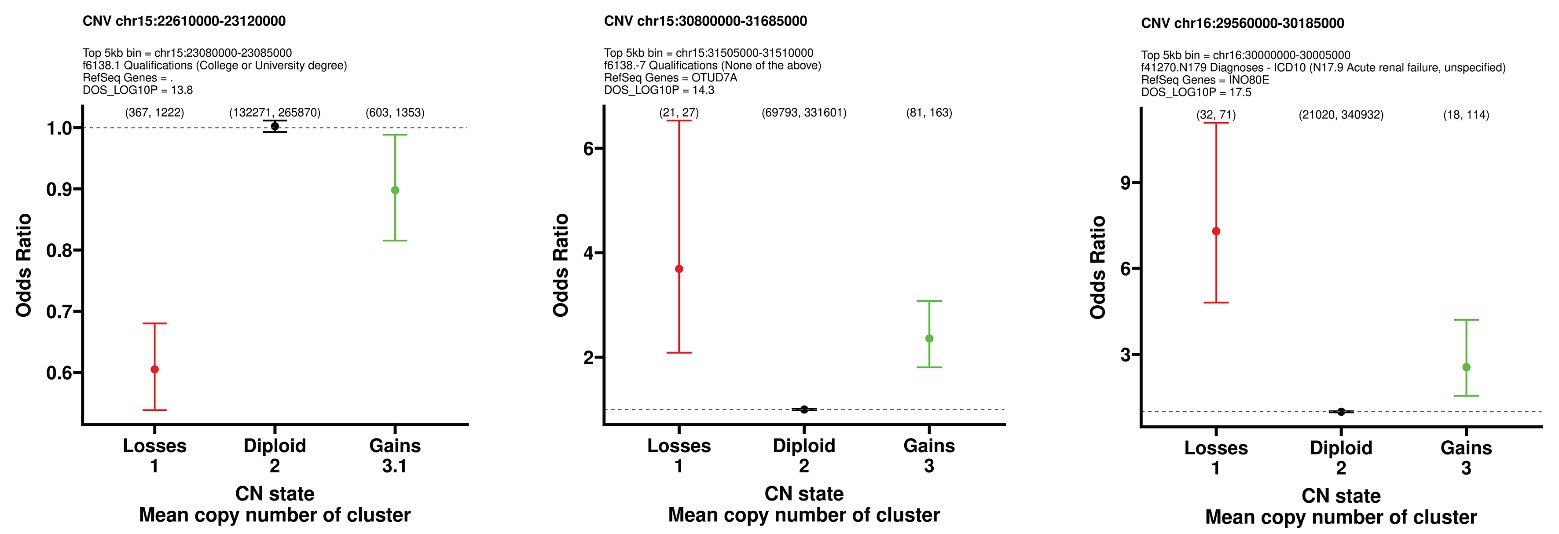

**Supplementary Figure 5. Data from three recurrent reciprocal deletion/duplication regions (15q11.2, 15q13.3 and 16p11.2) indicates that deletion of dosage sensitive genes within these regions typically has more severe effects than the reciprocal duplication.** Each plot shows odds ratio (y-axis) versus copy number state for the 5 kb bin with the strongest association within each recurrent reciprocal CNV region. Vertical bars represent 95% confidence intervals, with numbers in parentheses indicating the number of individuals per copy number state with and without the trait.

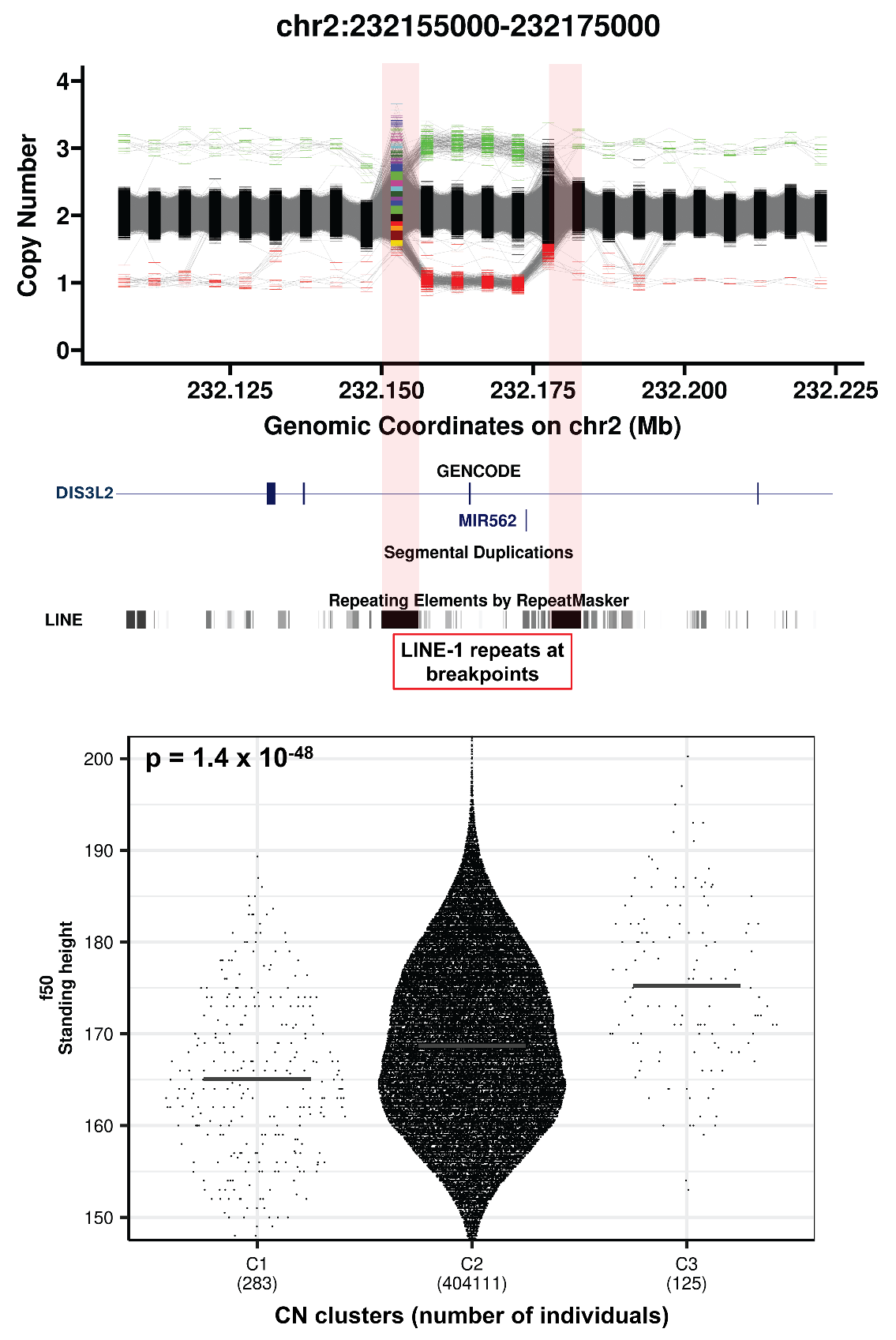

**Supplementary Figure 6. Copy number of a reciprocal gain/loss within *DIS3L2* is associated with standing height.** Upper plot shows CN estimates per 5 kb bin within *DIS3L2* within 2q37.1. Lower plot shows standing height (y-axis) versus copy number state for the 5 kb bin with the strongest association (chr2:232,155,000-232,175,000). In the upper plot, read shaded regions highlight a pair of LINE-1 repeats that occur at both breakpoints. In the lower plot, horizontal black bars show the mean of each distribution.

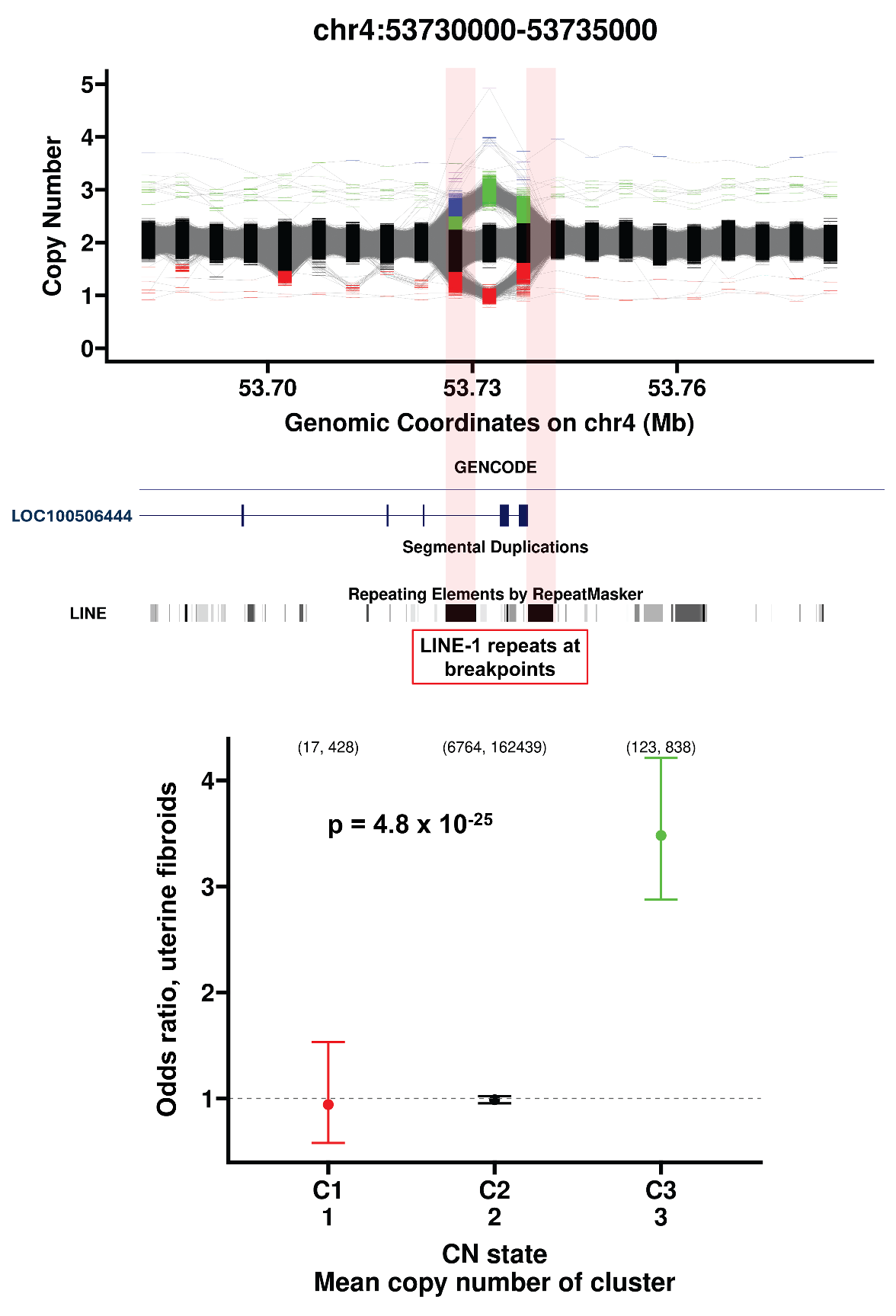

**Supplementary Figure 7. Copy number of a region overlapping the final exon of *LOC100506444* is associated with risk of uterine fibroids**. Upper plot shows CN estimates per 5 kb bin across the *LOC100506444* locus within 4q12. Lower plot shows odds ratio for uterine fibroids (y-axis) versus copy number state for the 5 kb bin with the strongest association (chr4:53,730,000-53,735,000). In the upper plot, read shaded regions highlight a pair of LINE-1 repeats that occur at both breakpoints. In the lower plot, vertical bars represent 95% confidence intervals, with numbers in parentheses indicating the number of individuals per copy number state with and without the trait.

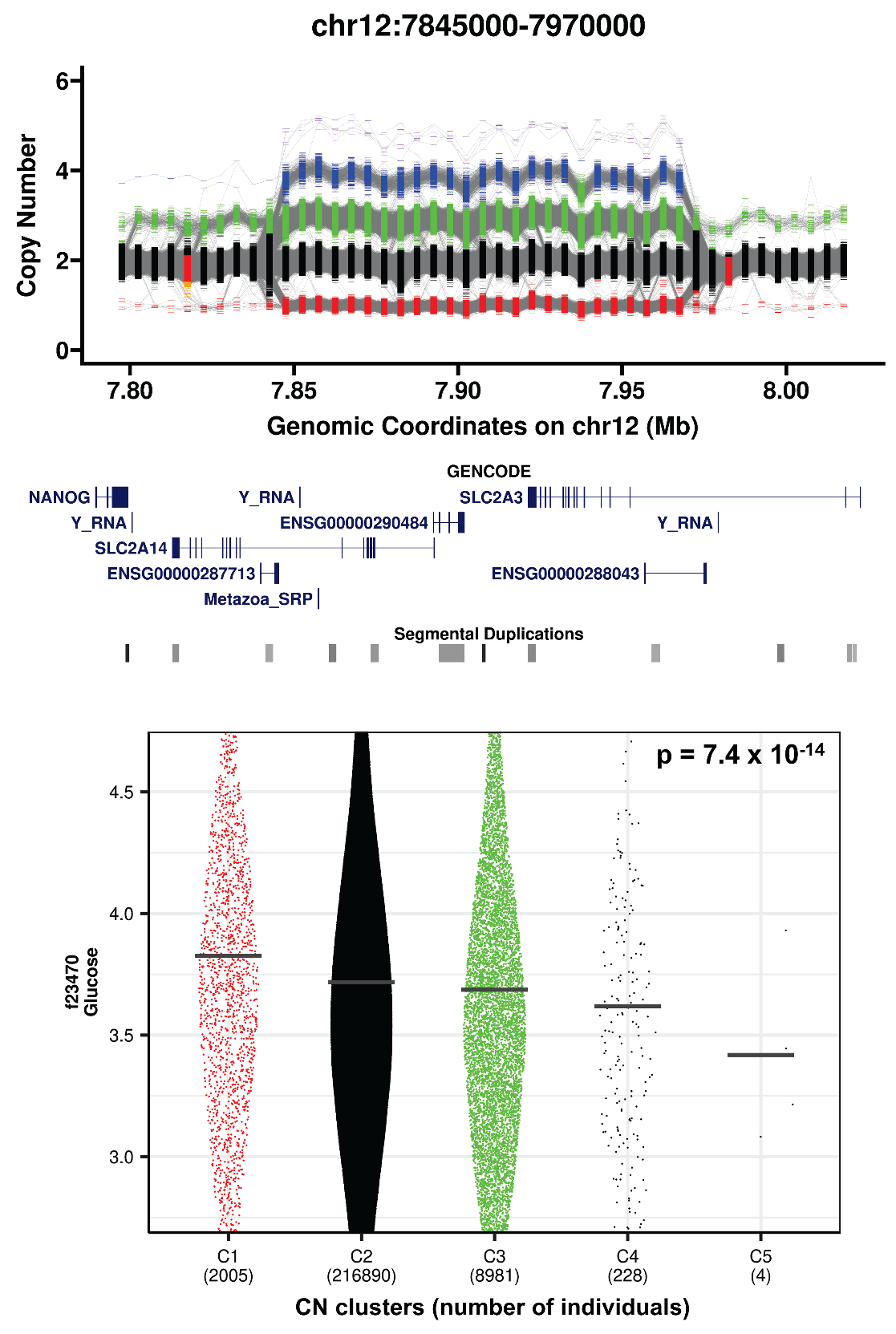

**Supplementary Figure 8. Copy number of a reciprocal gain/loss CNV that includes glucose transporters is associated with serum glucose.** Upper plot shows CN estimates per 5 kb bin within the *SLC2A3/SLC2A14* locus at 12p13.31. Lower plot shows serum glucose (y-axis) versus copy number state for the 5 kb bin with the strongest association (chr12:7,945,000-7,950,000). In the lower plot, horizontal black bars show the mean of each distribution.

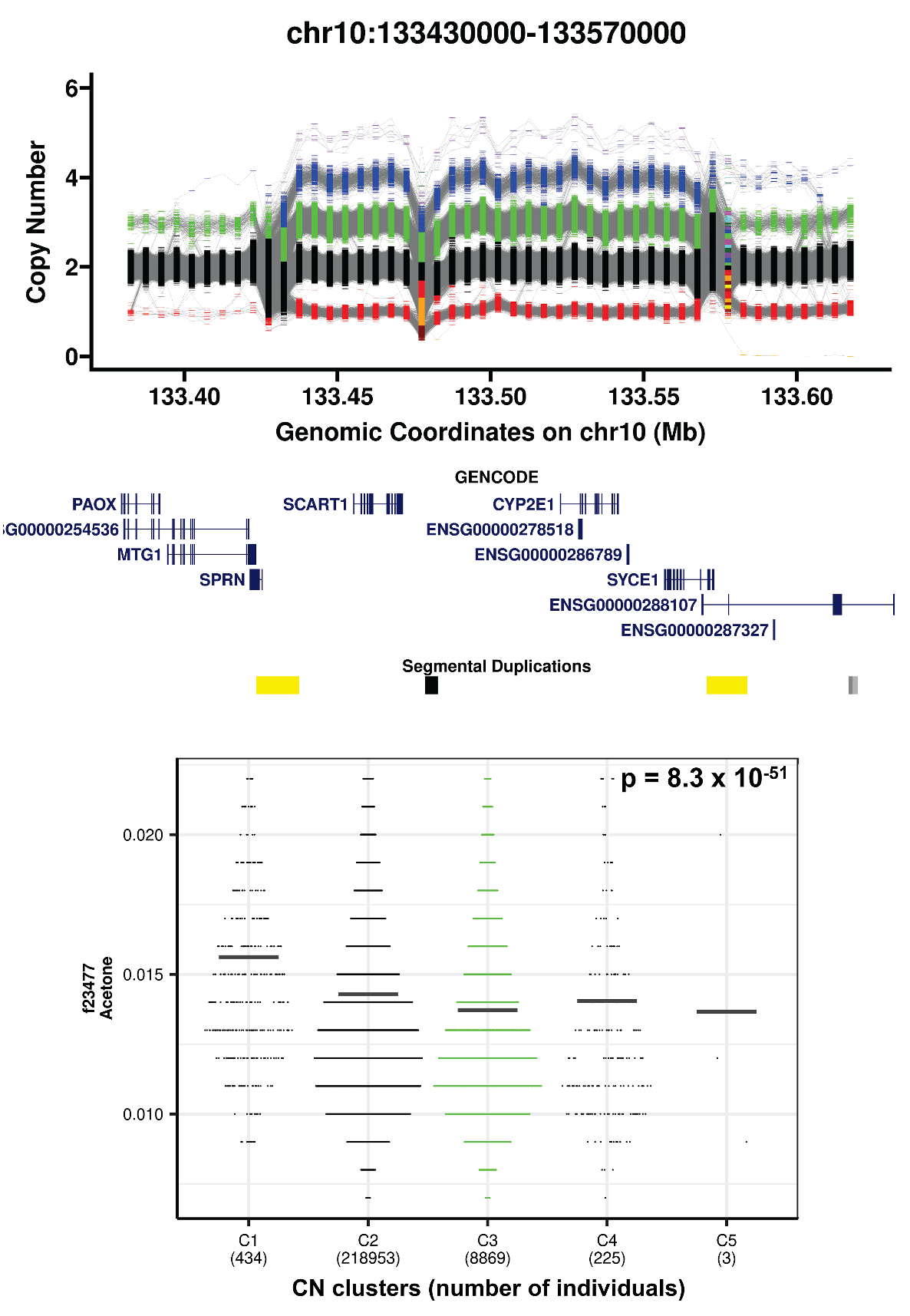

**Supplementary Figure 9. Copy number of a reciprocal gain/loss that includes *CYP2E1* is associated with serum acetone**. Upper plot shows CN estimates per 5 kb bin within the *CYP2E1* locus at 10q26.3. Lower plot shows serum acetone (y-axis) versus copy number state for the 5 kb bin with the strongest association (chr10:133,485,000-133,490,000). In the lower plot, horizontal black bars show the mean of each distribution.

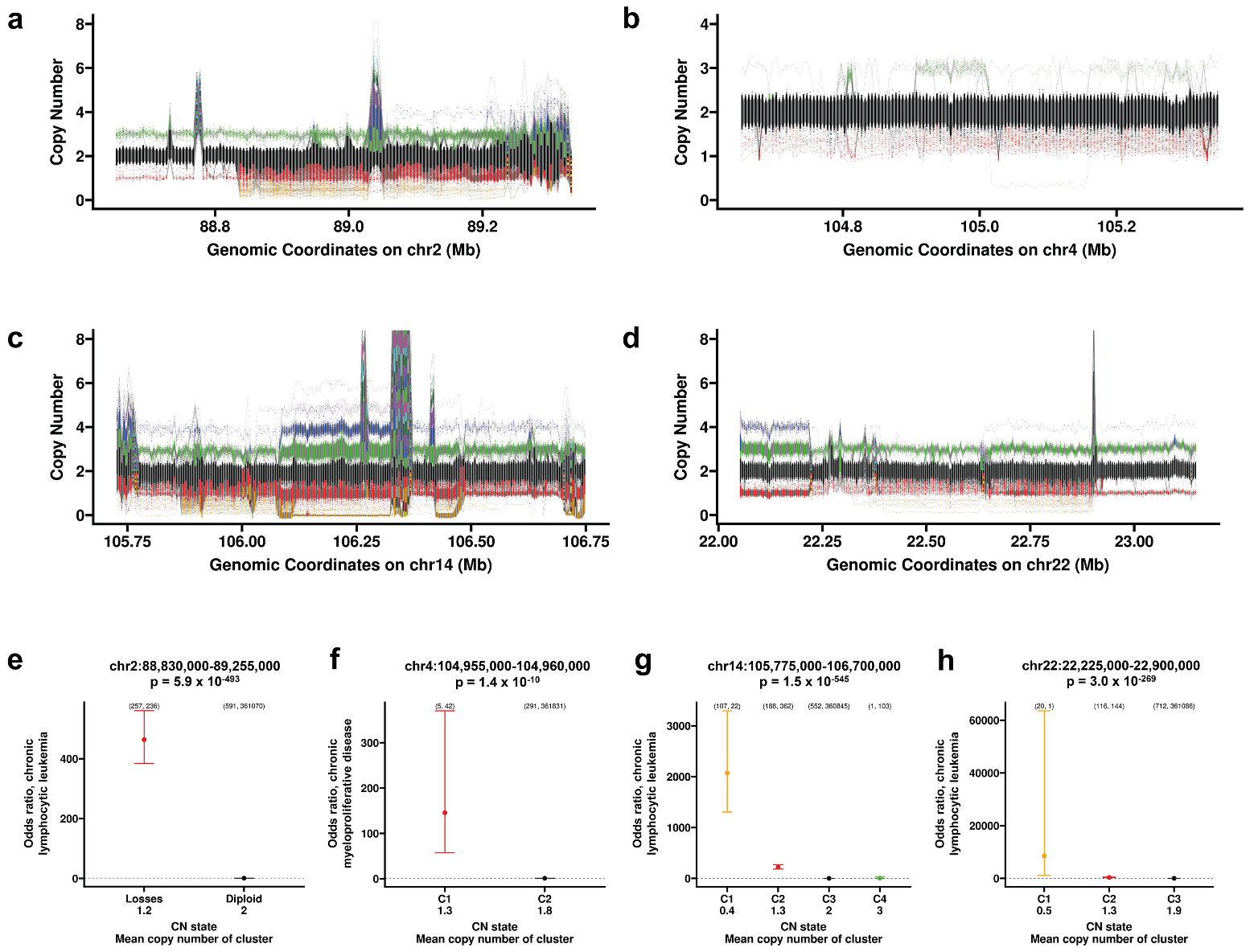

**Supplementary Figure 10. Mosaic deletions associated with high risk of leukemia.** (**a-d**) CN estimates per 5 kb bin within (**a**) the *IGKV* gene cluster at 2p11.2, (**b**) the *TET2* locus at 4q24, (**c**) the *IGHV* gene cluster at 14q32.33 and (**d**) the *IGLV* gene cluster 22q11.22. Lower plots show risk either chronic lymphocytic leukemia (**e, g, h**) or chronic myeloproliferative disease (**f**) versus copy number state for the 5 kb bins with the strongest association. In (**e-h**), vertical bars represent 95% confidence intervals, with numbers in parentheses indicating the number of individuals per copy number state with and without the trait.

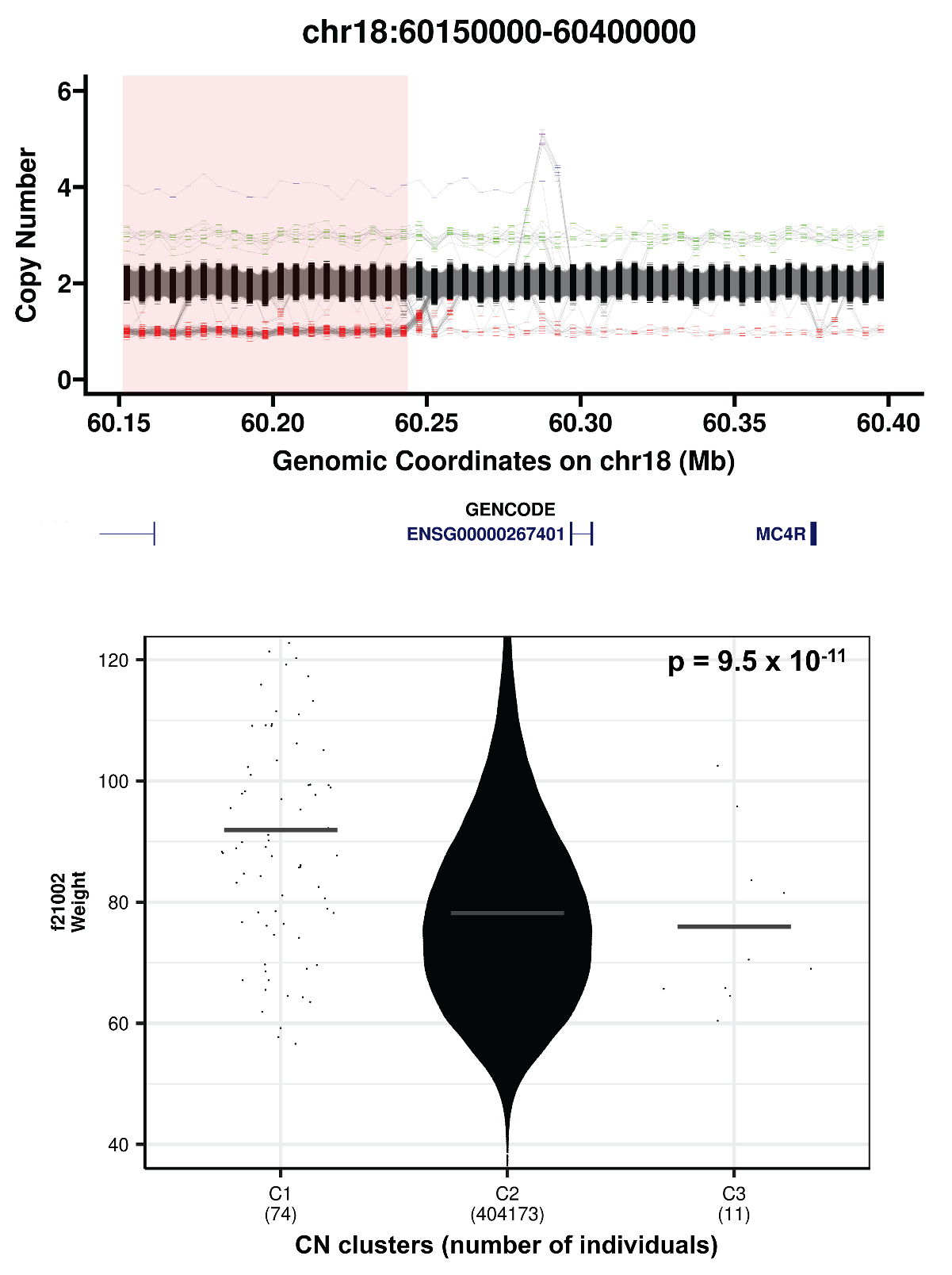

**Supplementary Figure 11. Deletion of a non-coding region ~100kb upstream of *MC4R* is associated with increased weight.** Upper plot shows CN estimates per 5 kb bin within the *MC4R* locus at 18q21.32. Lower plot shows weight (y-axis) versus copy number state for the 5 kb bin with the strongest association (chr18:60,210,000-60,215,000). In the upper plot, red shading indicates the deletion significantly associated with weight. In the lower plot, horizontal bars show the mean of each distribution.

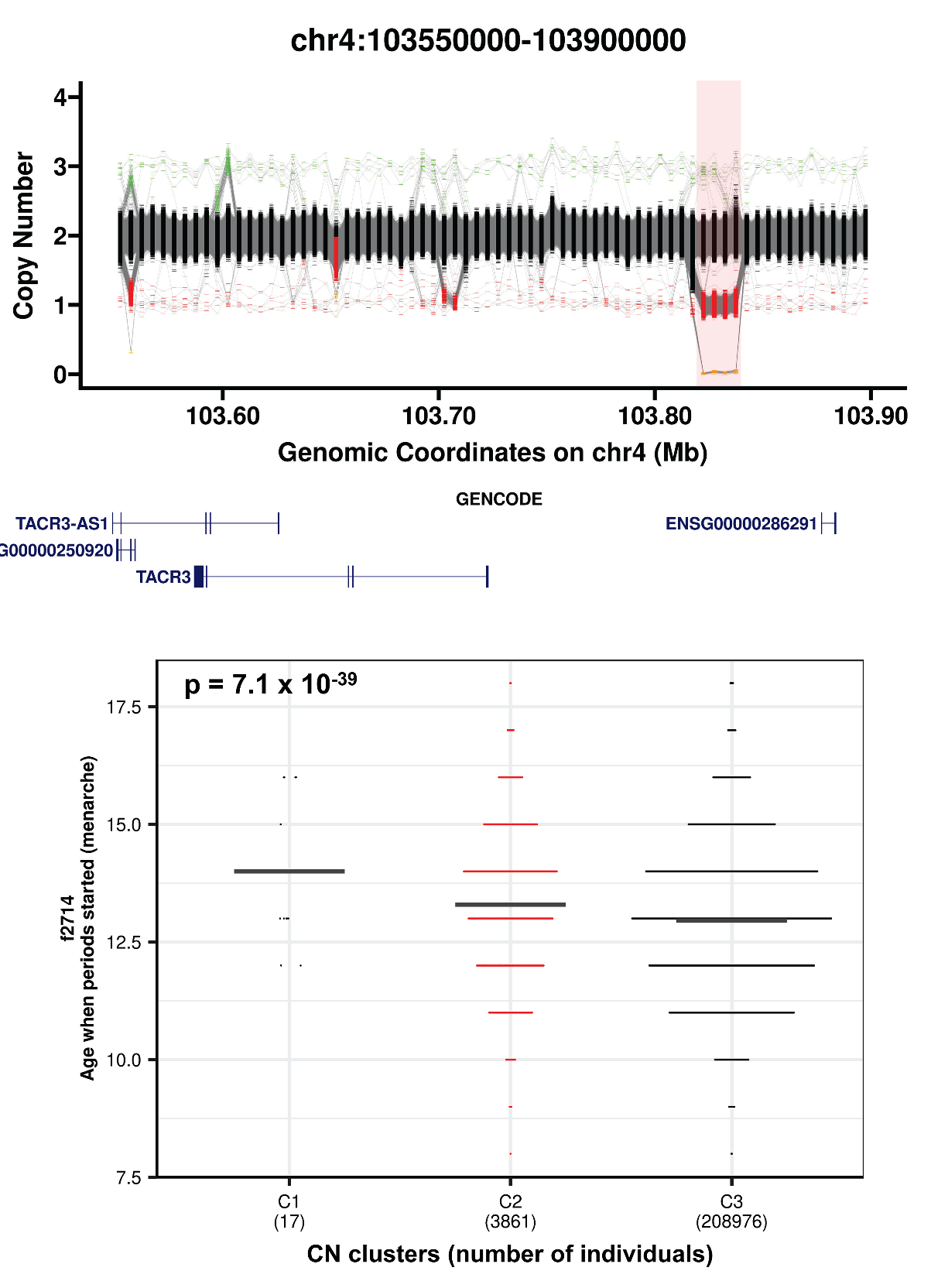

**Supplementary Figure 12. Deletion of a non-coding region ~100kb upstream of *TACR3* is associated with delayed age of menarche.** Upper plot shows CN estimates per 5 kb bin within the *TACR3* locus at 4q24. Lower plot shows age of menarche (y-axis) versus copy number state for the 5 kb bin with the strongest association (chr4:103,820,000-103,825,000). In the upper plot, red shading indicates the deletion significantly associated with age of menarche. In the lower plot, horizontal black bars show the mean of each distribution.

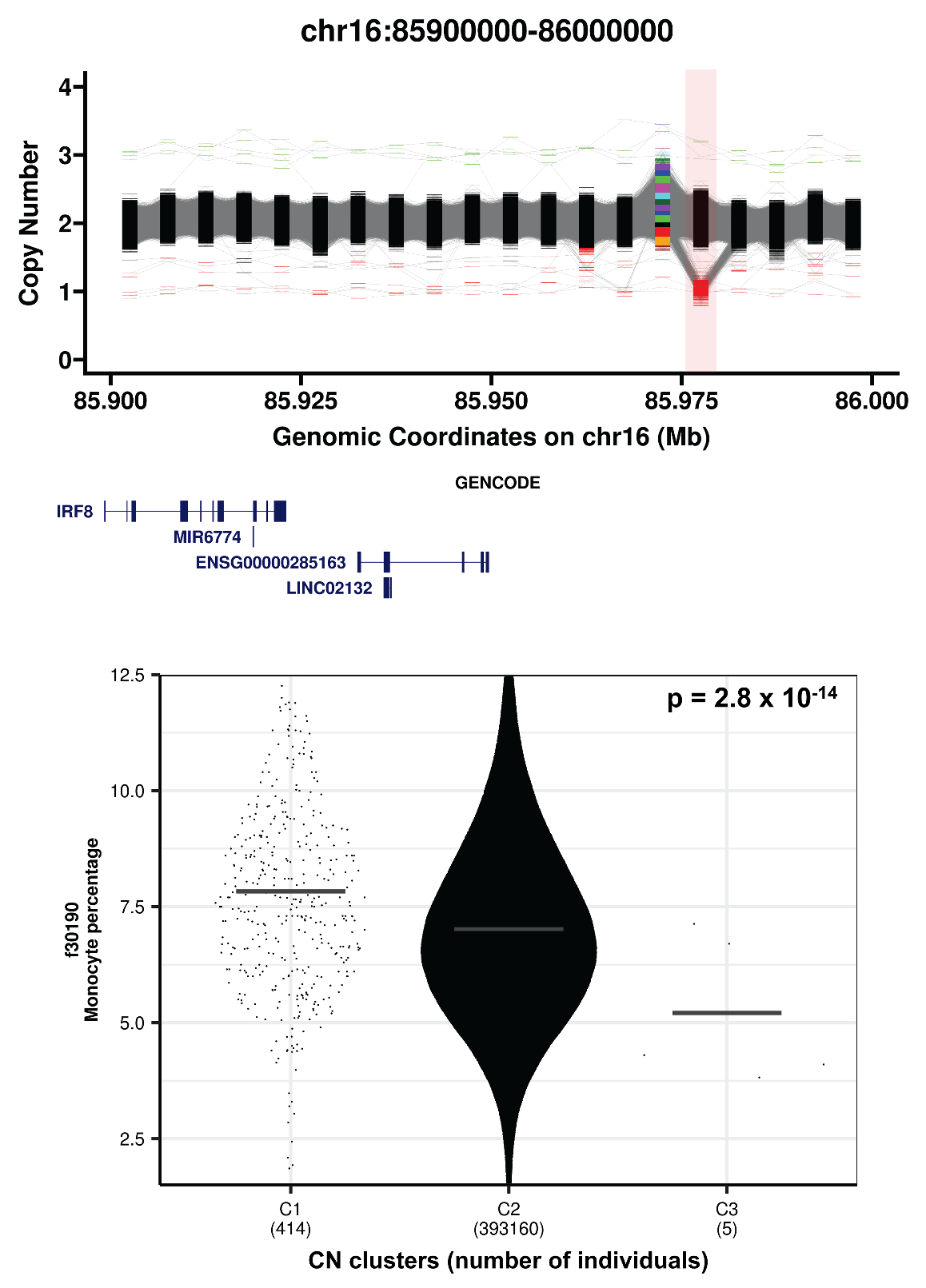

**Supplementary Figure 13. Copy number of a non-coding region ~70kb downstream of *IRF8* is associated with monocyte percentage.** Upper plot shows CN estimates per 5 kb bin within the *IRF8* locus at 16q24.1. Lower plot shows monocyte percentage (y-axis) versus copy number state for the 5 kb bin with the strongest association (chr16:85,975,000-85,980,000). In the upper plot, red shading indicates the deletion significantly associated with monocyte percentage. In the lower plot, horizontal bars show the mean of each distribution.

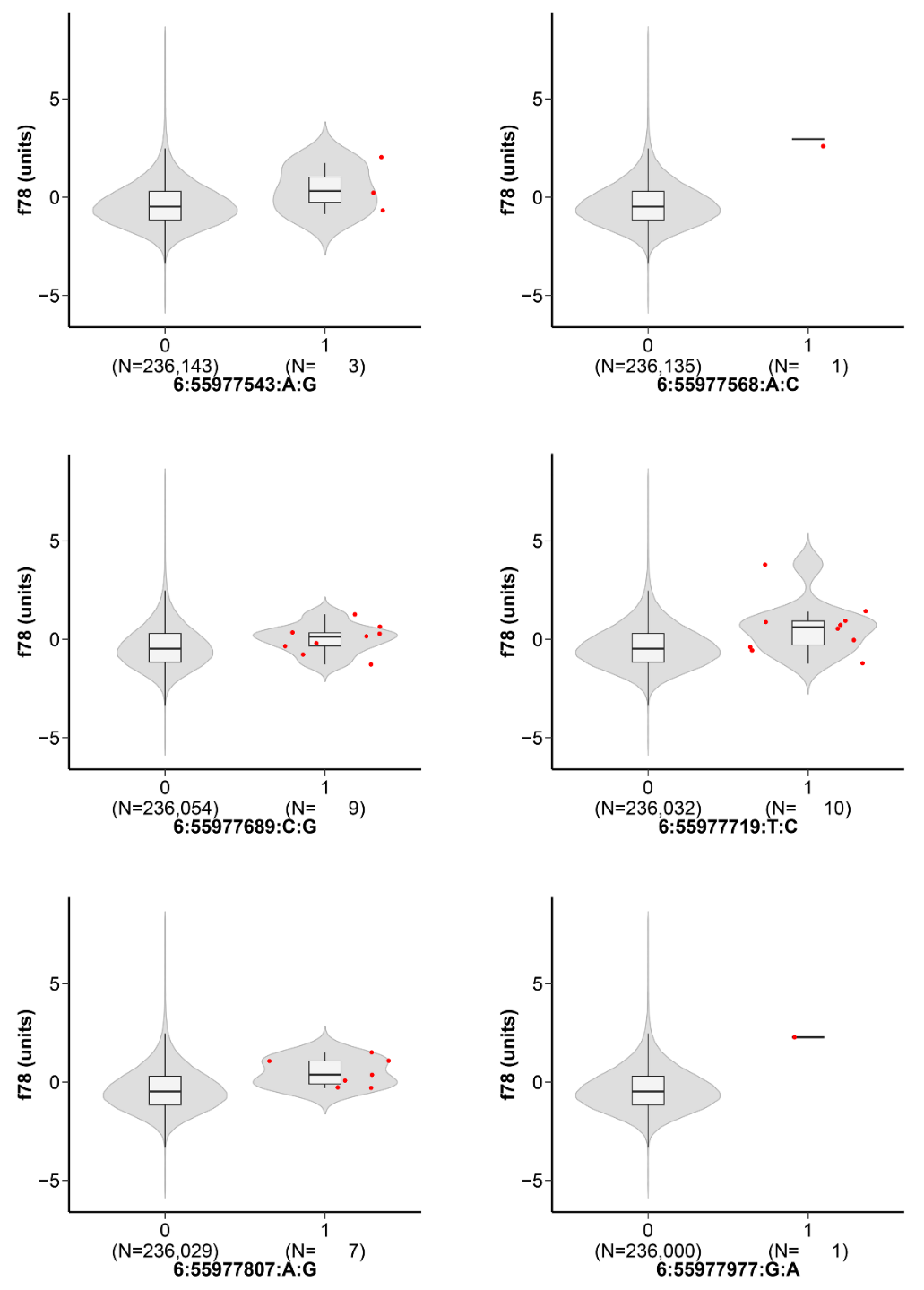

**Supplementary Figure 14. A cluster of rare constrained SNVs located within an annotated cis-regulatory element that lies within a non-coding deletion region that associates with bone mineral density also associate with the same trait and directionality as seen in deletion carriers.** Each plot shows data for one rare (MAF <0.001) constrained SNV, with bone mineral density t-score (y-axis) shown for individuals homozygous for the reference allele (“0”) versus those who carry the SNV in heterozygous form (“1”) (x-axis). Notation below each plot separated by colons indicate “chr:nucleotide position:reference base:alternate base” for each SNV. Within each violin, box limits indicate the 25^th^ and 75^th^ percentiles while the internal black bars show the medians; whiskers extend 1.5 times the interquartile range from the 25^th^ and 75^th^ percentiles. Red points show individual values where n < 50.

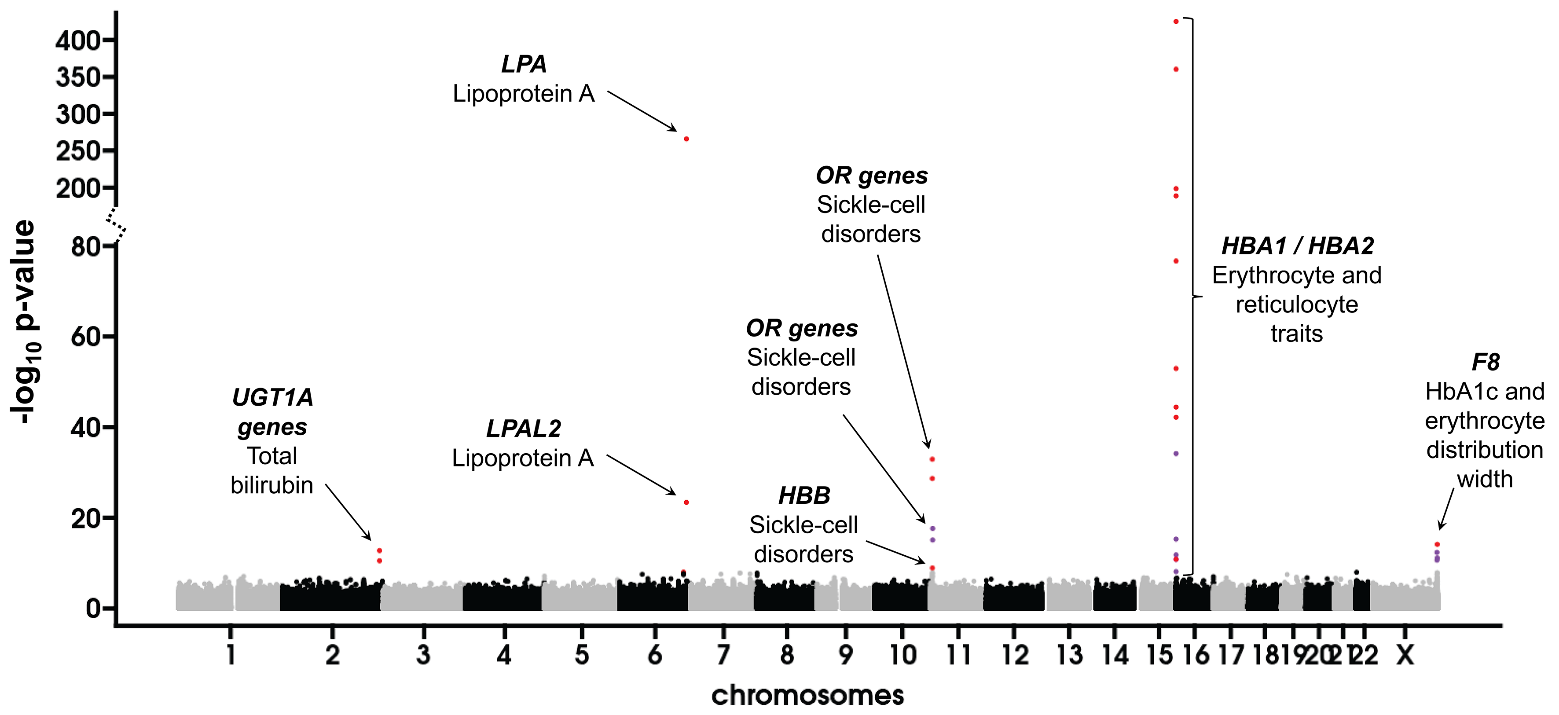

**Supplementary Figure 15. Manhattan plot showing associations identified by PheWAS of CNVs in individuals of African ancestry in the UKB cohort**. Points shown in color exceed the Bonferroni-corrected significance threshold of p < 9.1 x 10^-9^. Red dots indicate associations where the strongest signal was identified using an additive model, while purple dots represent associations where the strongest signal was identified using a dosage-sensitive model. Individual signals are annotated with gene name and trait. Note the discontinuous y-axis used due to the very strong signals observed at *LPA* and *HBA*1/2.

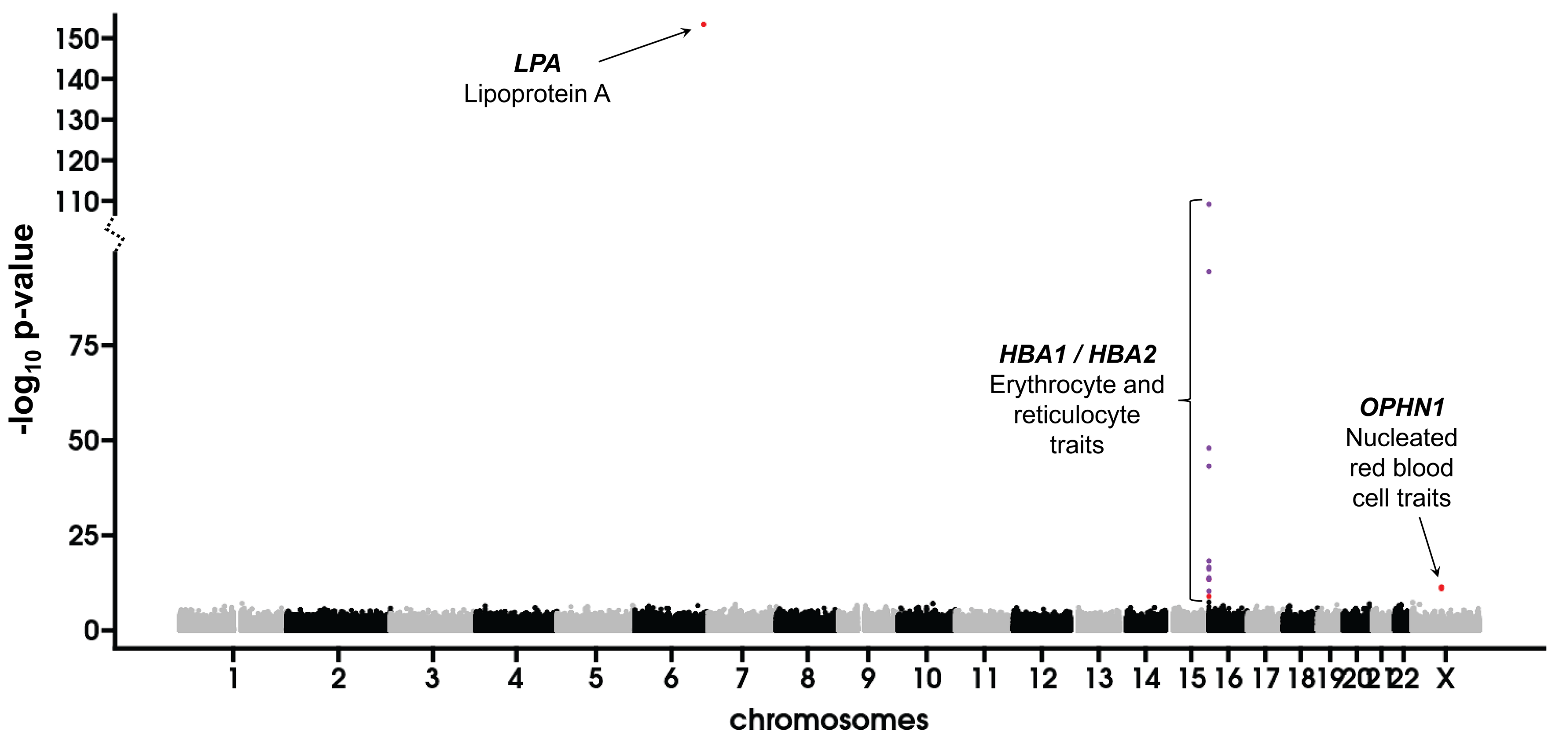

**Supplementary Figure 16. Manhattan plot showing associations identified by PheWAS of CNVs in individuals of South Asian ancestry in the UKB cohort**. Points shown in color exceed the Bonferroni-corrected significance threshold of p < 2.1 x 10^-8^. Red dots indicate associations where the strongest signal was identified using an additive model, while purple dots represent associations where the strongest signal was identified using a dosage-sensitive model. Individual signals are annotated with gene name and trait. Note the discontinuous y-axis used due to the very strong signals observed at *LPA*.

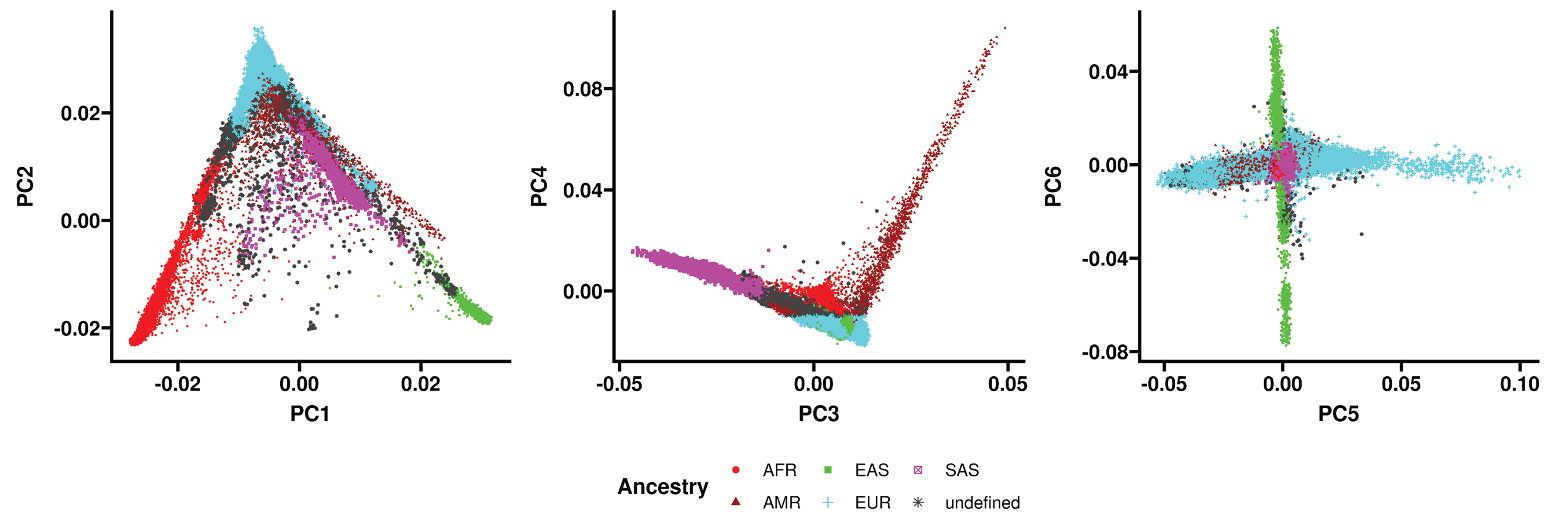

**Supplementary Figure 17. Ancestry prediction of UKB participants using principal component analysis ~65,000 high quality autosomal SNVs.** We retained samples predicted as either European, African or South Asian for PheWAS, representing the three largest ancestral groups present within the UKB cohort.

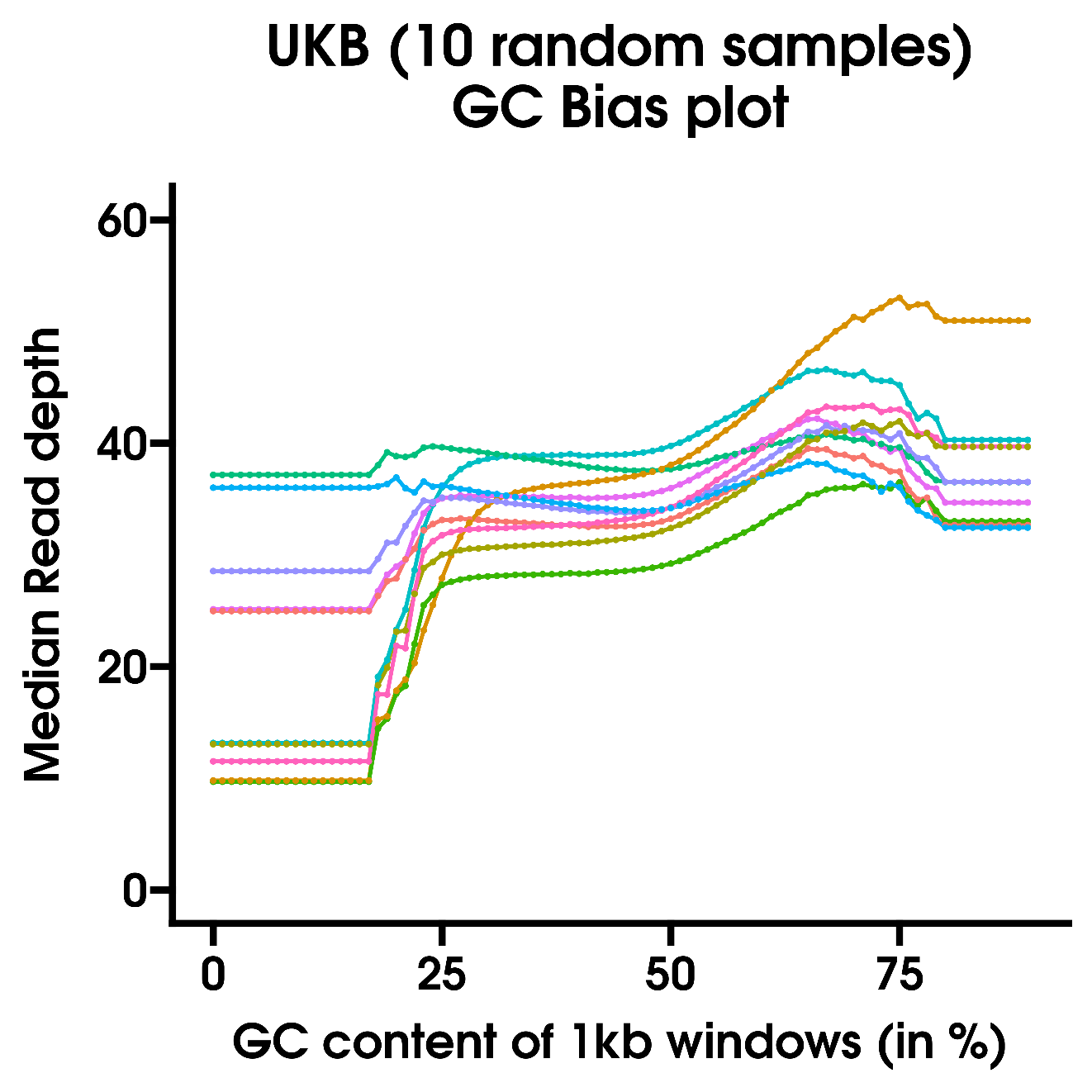

**Supplementary Figure 18. GC-bias varies among different sequenced samples.** The plot shows variations in median read depth among different samples based on GC-content, with each colored line corresponding to a different sequenced sample. We applied a correction factor per sample to normalize read depth for these variations.

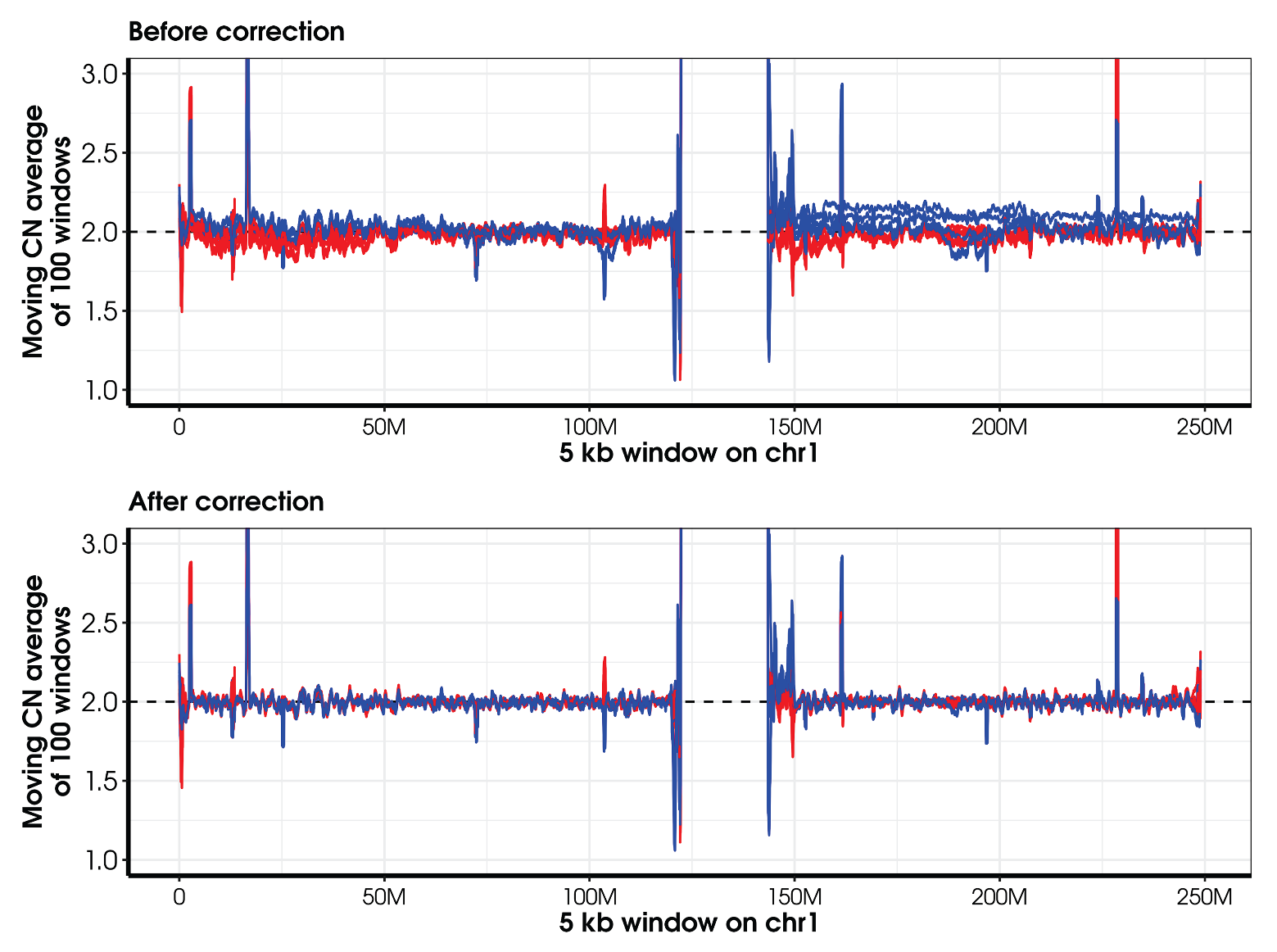

**Supplementary Figure 19. Correcting for “genomic waves” results in improved copy number prediction.** Genomic waves are systematic biases that can occur in copy number estimates from array and sequencing data. We calculated a genomic wave correction factor per 2 Mb bin per sample and used this to perform region-based normalization of copy number estimates in each individual. To illustrate the effect of this normalization, plots show copy number estimates before (top) and after (bottom) this correction in 10 samples that showed some of the highest mean deviations in copy number estimates on chromosome 1.

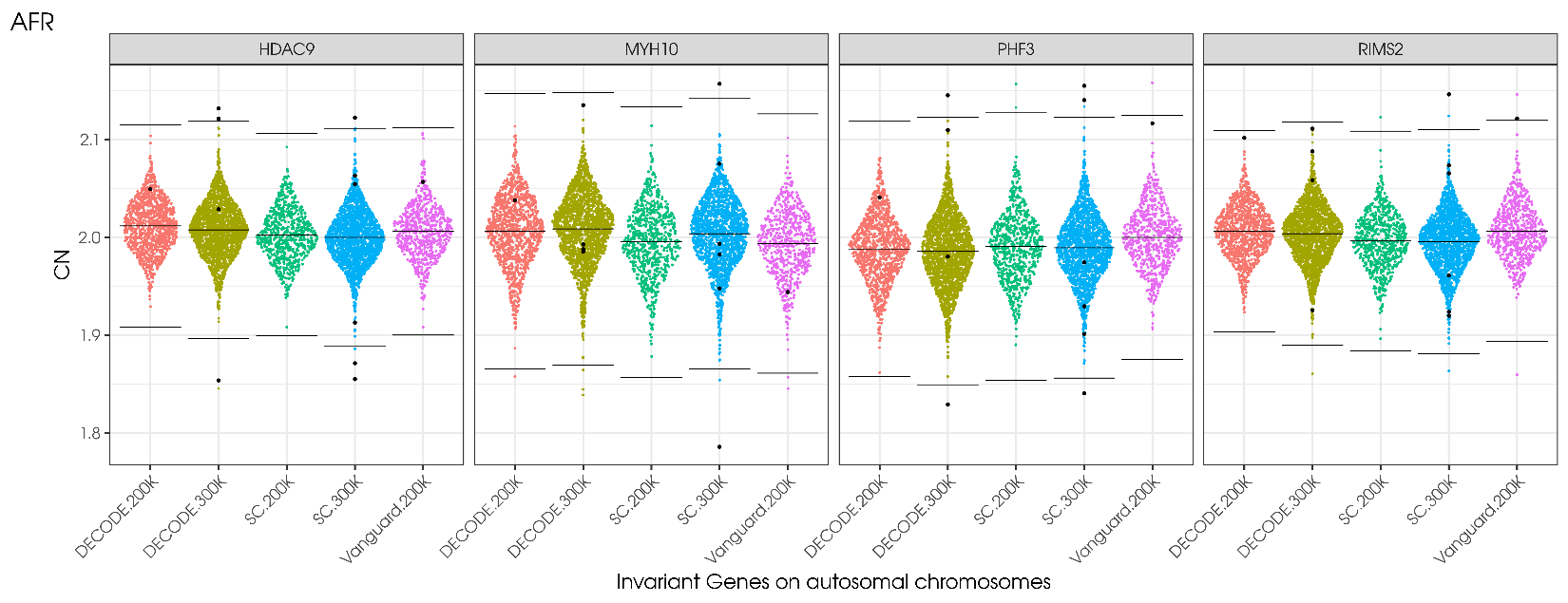

**Supplementary Figure 20. Removal of technical outliers and observation of systematic batch effects.** We calculated copy number estimates at 200 highly constrained autosomal genes, separating samples into the five major sequencing batches present in the UKB cohort. The plot shows data for individuals of African ancestry for four example genes used in this analysis. Systematic effects on copy number estimates among sequencing batches are evident, which we adjusted for by applying median correction to samples from each batch per 5kb window across the genome. In addition, samples shown as large black dots were consistent outliers for copy number estimates (≥4 standard deviations from the median at ≥10 highly constrained genes) and were removed from further analysis. For each distribution, horizontal black bars represent the median and ± 4 standard deviations.

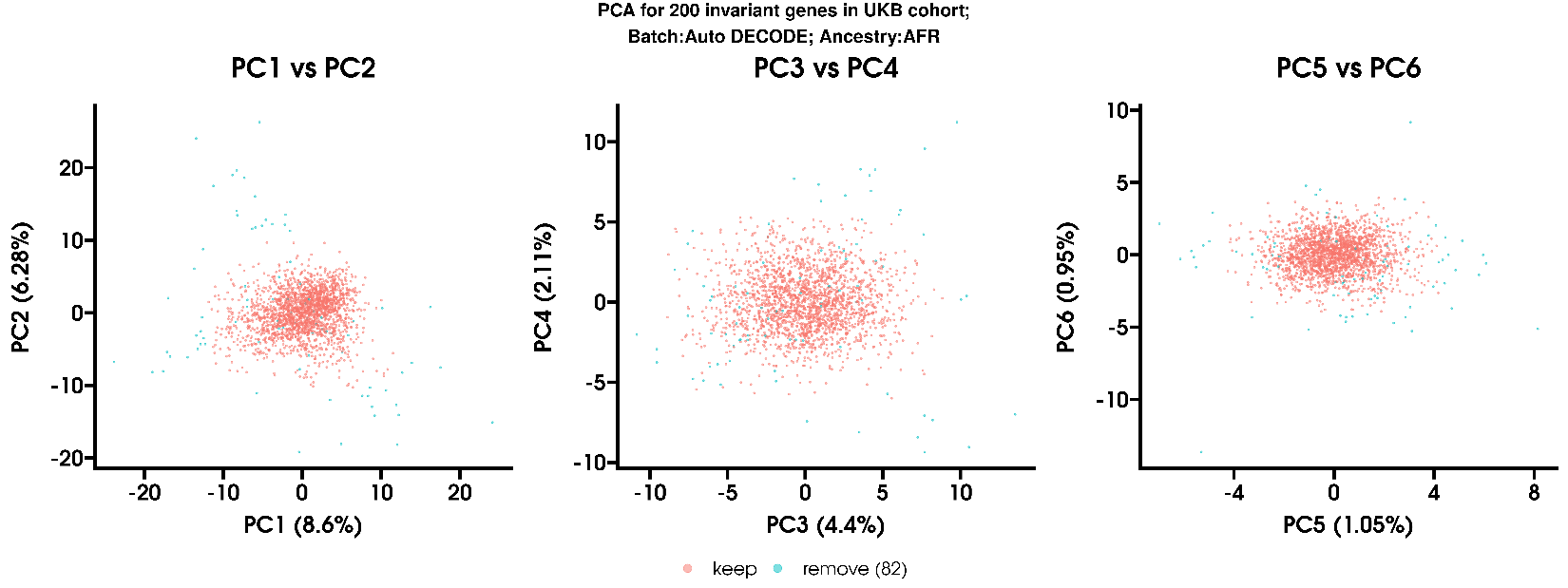

**Supplementary Figure 21. Identification of technical outliers for read depth using principal component analysis of copy number estimates at 200 highly constrained autosomal genes.** Plot shows data for individuals predicted to be of AFR ancestry. Points shown in blue were considered outliers and were removed from further analysis.

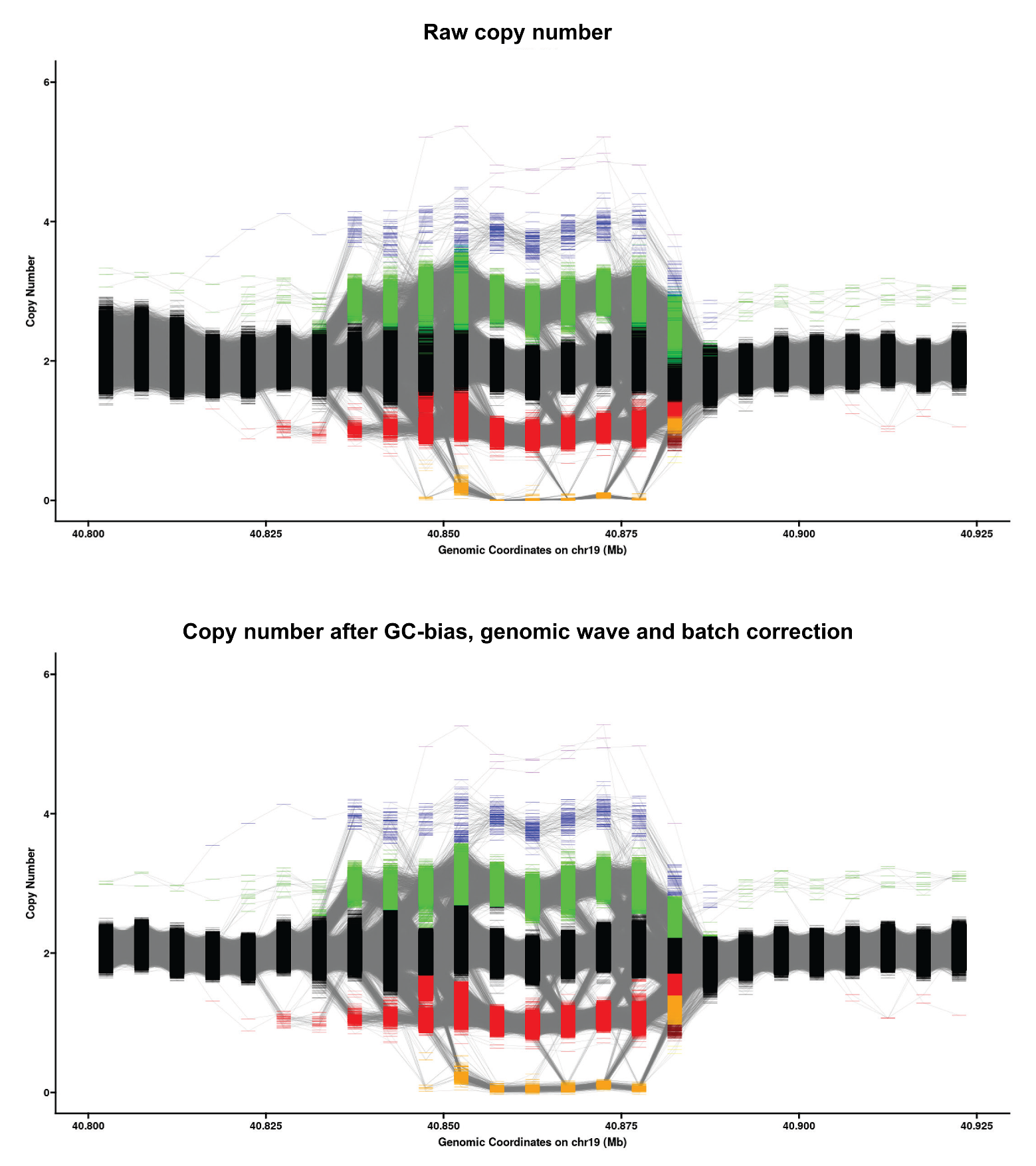

**Supplementary Figure 22. Estimated genomic copy number based on raw (pre-normalized) data and after correction for GC-bias, genomic wave and batch.** Following these normalization steps (lower plot), copy number estimates per 5kb bin are more consistent, showing tighter distributions and improved separation between genotype clusters. The improvement in data quality is most evident on the left side of the plot. Assigned genotype clusters are shown as different colored points.

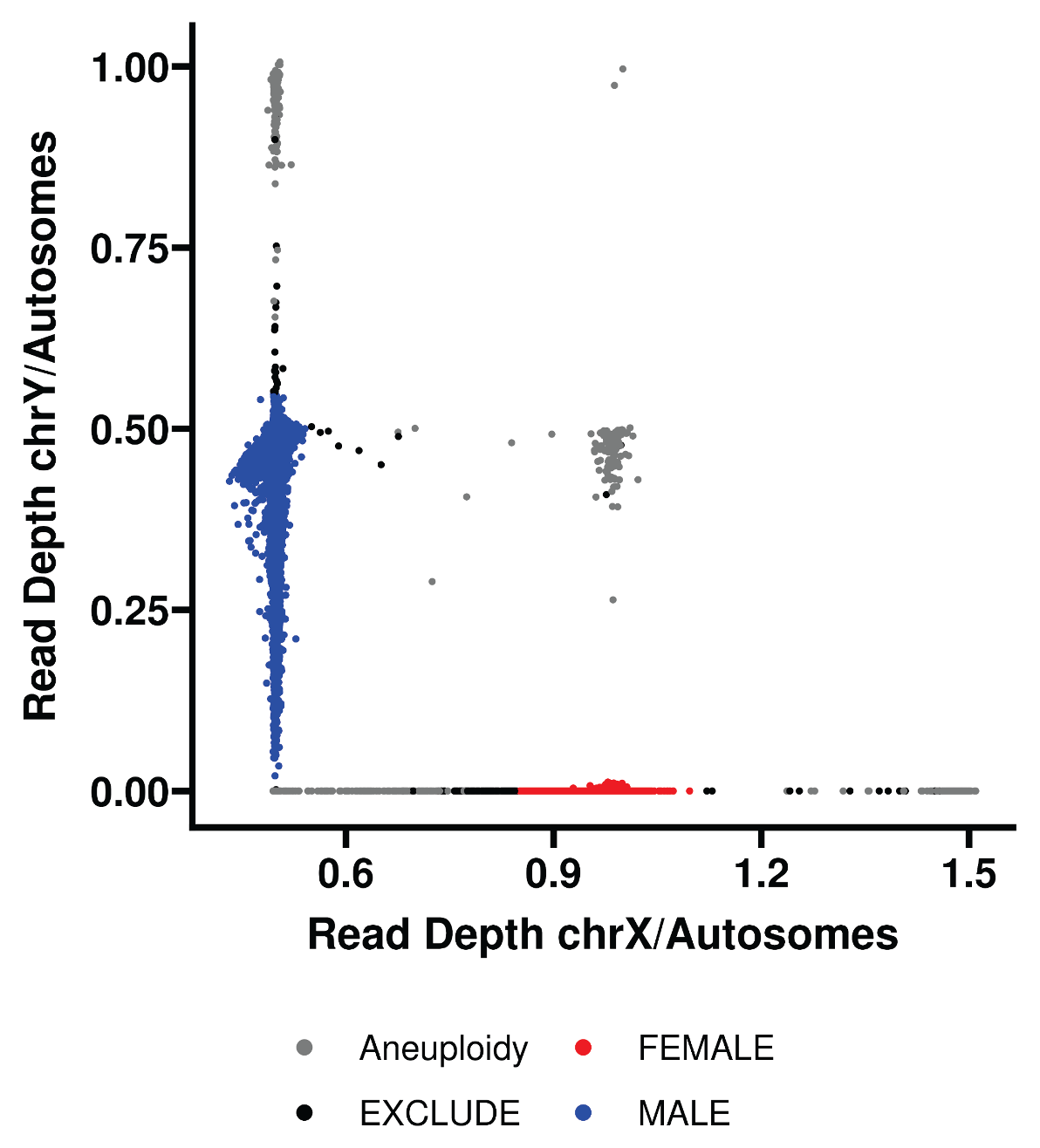

**Supplementary Figure 23. Determination of sex chromosome complement using the relative ratio of median read depth on the sex chromosomes compared to autosomes.** Samples with predicted sex chromosome aneuploidy or which were outliers from the main clusters were removed from further analysis.

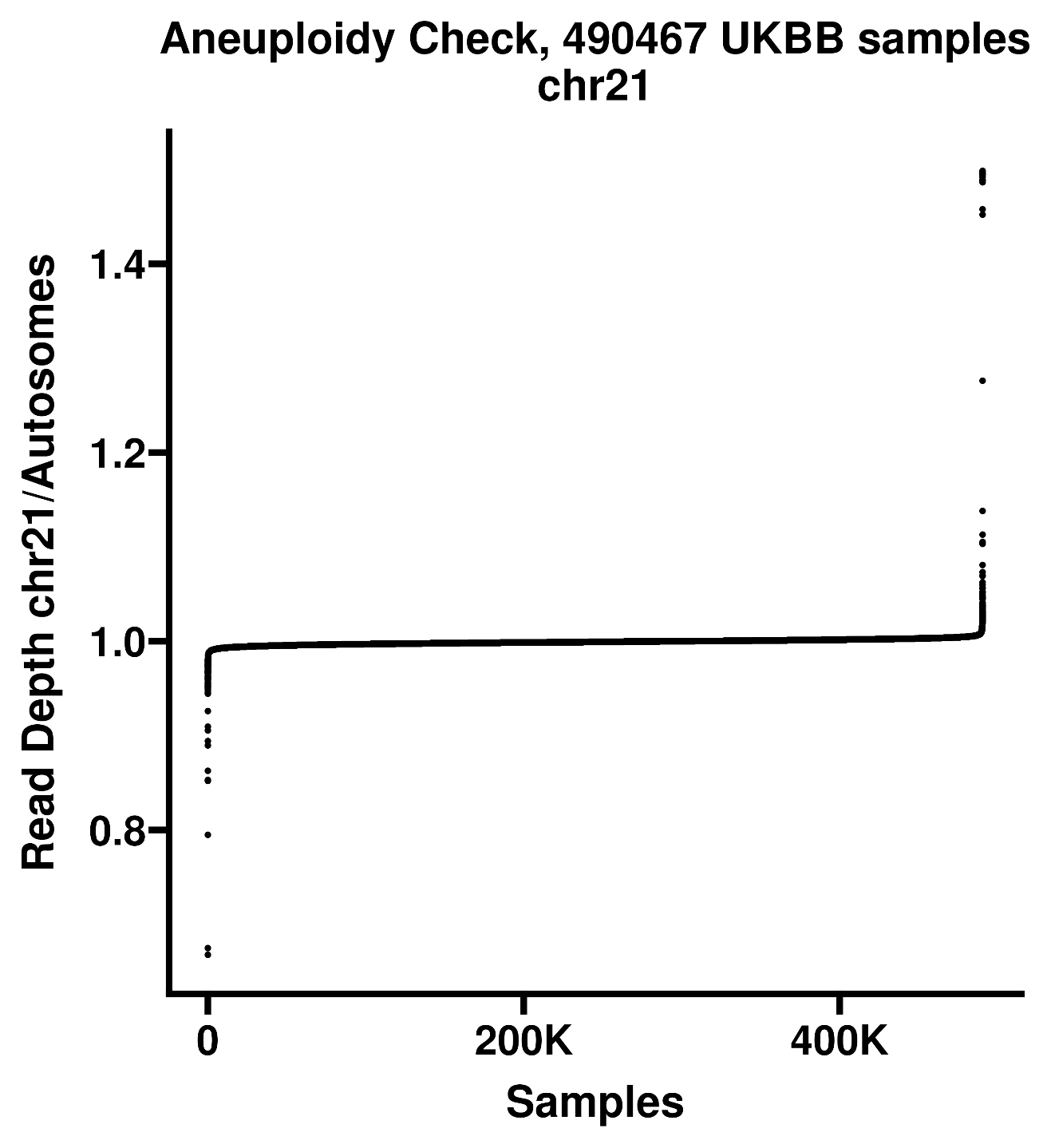

**Supplementary Figure 24. Determination of samples with autosomal aneuploidy.** We used the relative ratio of median read depth per chromosome compared to median read depth across chromosomes 1-22. The plot shows results for chromosome 21 with individuals sorted based on the ratio of relative median read depth on chromosome 21 to the autosomes. Five individuals are predicted to have complete trisomy 21 (top right corner of plot), with ratios of read depth on chromosome 21/autosomes of ~1.5, while additional individuals at each tail of the distribution are either technical outliers or have partial aneuploidy or mosaic monosomy / trisomy of chromosome 21 and were removed from further analysis. Similar analysis was performed for every autosomal chromosome.
